# _CLIA_MDK: A Modular Smartphone Platform Matching Plate Reader Performance for Chemiluminescent Immunoassay Development

**DOI:** 10.64898/2026.03.26.26348440

**Authors:** Christopher S. Wood, Seraina M. Abele, Jannis Alsbach, Ariola Gervalla, Dominik M. Meinel, Andreas P. Cuny

## Abstract

The development of chemiluminescent immunoassays (CLIAs) is a complex and iterative process that relies on costly laboratory infrastructure, limiting its accessibility and application across healthcare settings and disease areas. Here, we detail the CLIA Mobile Development Kit (_CLIA_MDK) a modular, mobile, and inexpensive platform to assess image sensors, smartphones and data processing workflows for CLIA development. For its demonstration, we developed two CLIAs targeting renin and aldosterone, key biomarkers for diagnosing primary aldosteronism. The results from our performance study, including 50 patient samples, demonstrate the potential of our platform in a real-world scenario. We found that the performance of our mobile reader platform is comparable to that of a state-of-the-art plate reader, with a Lower Limit-of-Detection (LLoD) approaching 41 femtomolar. We envision that our platform will help accelerate CLIA development, make it more accessible, and lay the foundations for novel, distributed, yet highly sensitive diagnostic tests.

## 1 Introduction

Chemiluminescence immunoassays (CLIAs) combine antigen-antibody specificity with sensitive chemiluminescent detection, enabling quantification of very low analyte concentrations in biological samples^1^. Widely adopted in clinical labs, CLIAs provide rapid, accurate results over a broad concentration range^2,3^. They include a chemiluminescent label, affinity reagents optimized for rapid binding, immobilization methods, and a detector. Commercial CLIA analyzers are often large, costly, and require dedicated lab space, limiting access^4^.

Vendors are reluctant to support tests for rare diseases due to low throughput and prevalence^5–7^, and “homebrew” assay transfers remain costly and constrain development flexibility through system lock-in^8^. In the spirit of open science, low-cost and open platforms are needed for co-developing assays and readout technologies, as current systems rely on costly, assay-specific instruments and lack pathways for prototyping^9,10^.

One contributor to the cost of the lab-based CLIA systems is the optoelectronic detector. Fortunately, over the last decade, the popularity of smartphones capable of taking photos in low light conditions has driven successive improvements in Complementary Metal-Oxide-Semiconductor (CMOS) image sensor quality and availability^11,12^. Recent smartphone models even feature large sensors (e.g., Sony IMX989 1 inch), comparable to mid-range SLR cameras in terms of image quality, sensitivity, and raw data linearity^13,14^. As the performance of new CMOS image sensors in smartphones has improved and the popularity of mass-manufactured smartphones has increased, the price of older sensors has decreased, increasing the availability of low-cost standalone camera modules, such as camera serial interface (CSI) cameras or embedded cameras^15,16^.

CMOS image sensors, particularly those integrated into smartphones, play an increasing role in mobile health applications and increase connectivity to the internet and access to diagnostics in low and middle-income countries^17,18^. These sensors have seen increasing application in luminescent assay readouts^19,20^, with a growing emphasis on point-of-care diagnostics^10,21^. They have also shown potential in various biomedical and clinical domains, including infectious disease detection^22–30^, medical screening^31,32^, as mobile microbiological laboratories^33^, and advanced imaging^34–36^. Despite this progress, there remains a critical need for a cost-effective (<$100), modular platform to systematically evaluate how sensor hardware and parameters affect assay performance^9,37^. An example demonstrating this need is the diagnosis of endocrine disorders such as primary aldosteronism (PA), where accurate point-of-care measurement of the key biomarkers renin and aldosterone is clinically important but currently unfulfilled^38^. Accurate renin detection in blood samples is particularly technically challenging due to its low abundance and the confounding cryoactivation effect of prorenin conversion during sample freeze–thaw cycles^39,40^.

A modular, mobile evaluation platform that can benchmark sensor and assay performance against state-of-the-art (SOTA) laboratory instrumentation, including luminescence-capable plate readers, is essential to facilitate assay development and validation, and address these challenges.

Here, we introduce a 3D-printed modular, small-footprint system designed for assessment of various image detectors — including smartphone camera sensors and CSI camera modules — under controlled acquisition parameters (e.g., exposure time, sensitivity, aperture) and post-processing methods *in silico* and *in vitro*. We demonstrate the utility of the (ChemiLuminescent ImunnoAssay Mobile Development Kit _CLIA_MDK) platform using two self-developed CLIAs targeting renin and aldosterone. Our platform enables performance comparison with commercial laboratory plate readers and FDA-approved reference assays. When combined with a smartphone, our platform demonstrated comparable analytical performance to the SOTA and is a low-cost alternative for accurate biomarker detection, with an LLoD approaching 41 femtomolar. In summary, this work demonstrates a platform that will significantly accelerate CLIA-based diagnostic development and increase access to this highly sensitive assay technology worldwide.

## 2 Results

### 2.1 Quantitative image sensor evaluation method for the development of chemiluminescent immunoassays

To exploit widely available image sensors present in smartphones and facilitate their use in the development of low-cost CLIAs, we developed a multi-stage modular platform (**Fig. 1a**). In the first stage, the Mobile Camera Characterization System (MCCS) allows for the rapid and low-cost *in silico* evaluation of different smartphone image cameras, standalone CSI camera modules, and associated image sensor parameters (**Fig. 1a,b**). In the second stage, the mobile reader (MR) allows testing of the candidate sensors under assay conditions contained in standard 8-well strip labware. The compatibility with standard microplate formats means seamless integration with laboratory automation systems, and that the assays can be read with SOTA plate readers (PR) (**Fig. 1a,c**). In the last stage, relevant clinical ranges are evaluated with standard curves following established FDA guidelines^41,42^ by the computation of limit of quantification (LoQ), lower limit of detection (LLoD), and upper limit of detection (ULoD) (**Fig. 1a,c**). When the relevant clinical range is covered, a performance study on patient samples can be initiated. Otherwise, if the detection limits fall within the clinical range, an optimization cycle is started to optimize acquisition parameters, enhance signal processing, or adjust assay parameters until the requirement is met. This process is guided by our generic mobile app, which enables the wireless control of the MCCS’s light-emitting diodes (LEDs), including their color and brightness via an 8-bit input range, and control over the image acquisition mode and image sensor parameters (**Fig. 1d, Suppl. Fig. 1a-d**). For the acquisition of a typical assay standard curve, we perform the assay on blank samples and a set of concentrations of the target protein in pooled human serum matrix, referred to as spiked samples (**Fig. 1e**). Then, an image is acquired and processed to automatically detect the reaction wells and quantify the signal within a square region of interest (ROI) within the well using a modified version of our previously developed software pyPOCQuant^43^. In brief, using the well center and a mean signal accounts for the potential inhomogeneous distribution of the CLIA emission, such as at the well borders (**Fig. 1e,f**). We found that the side length variations of the ROI have a negligible effect on the signal distribution and the mean **Suppl. Fig. 2a-f**. For our experiments, we use a ROI side length of 80 pixels for the MCCS experiments and 60 pixels for the assay experiments, unless stated otherwise. We quantified the mean raw pixel values in the ROI (termed signal hereafter, see **Methods**) per primary color channel (RGB) to determine the optimal primary image channel for quantification of the chemiluminescent emission for increased sensitivity or to account for potential image sensor saturation (e.g., for 5000 pg mL^−1^ in channel blue **Fig. 1f**). Signals are associated with sample concentrations in each well to generate a quality control plot, including significance tests to ensure that the various concentrations can be separated, thereby helping to identify relevant parameters (**Fig. 1a,f**.)

**Figure 1:**
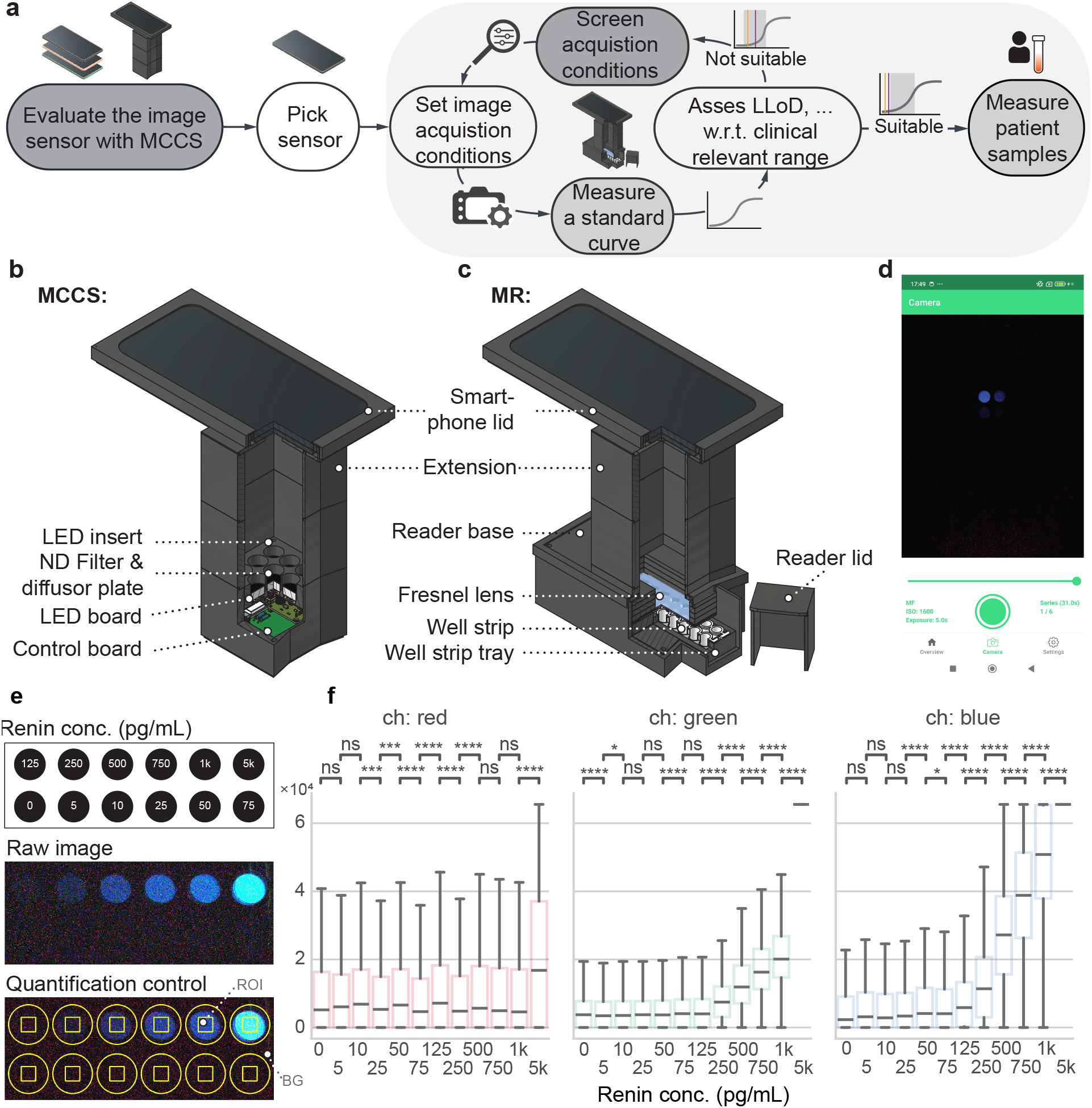
Mobile platform for the evaluation and development of CLIAs. **a** Workflow of the development and evaluation platform, including the selection of an image sensor, its acquisition conditions, measuring standard curves, and optimization studies. **b** Schematic illustration of the Mobile Camera Characterization System (MCCS) for the *in silico* sensor and signal processing evaluation. **c** Schematic illustration of the Mobile Reader (MR) for CLIA signal quantification. **d** Mobile app to control the MCCS and MR. **e** Quantification control and analysis for a biomarker standard curve targeting Renin. Analyte concentrations, the resulting raw image, and control of well detection accuracy (yellow circle). A squared region of interest (ROI, yellow box) and background (BG) indicate some of the areas assessed for signal and background quantification, respectively. **f** Box plot of raw pixel values from ROI as a function of the biomarker concentration analyzed by primary image channel (red, green, blue) with pairwise statistical significance test. The box represents the inter-quartile range (IQR), with the median (hereafter termed signal) shown as a horizontal line. Whiskers extend to the most extreme data points within 1.5 times the IQR. n = 3721 pixels.

### 2.2 Choosing the most suitable mobile sensor, parameters and pre-processing *in silico*

To efficiently assess a new camera sensor’s performance in quantifying CLIA signals and reduce use of valuable samples and reagents, we established an *in silico* workflow enabling pre-experimental condition screening (**Fig. 2a**). The MCCS and custom app enable testing different sensors, parameters, and signal enhancement algorithms to identify optimal configurations. We evaluated smartphone cameras (Xiaomi 13 Pro, Huawei P60 Pro, P20, Fairphone 5, Nokia G22) and CSI cameras (Arducam 3MP, 5MP, OmniVision 5640) for low-cost, high-sensitivity CLIA applications. First, we determined the sensor’s limits of detection, specifically the lowest brightness detectable (**Methods**). Among tested cameras, the X13P showed the highest sensitivity, handling the most neutral density (ND) filters while detecting a signal **Fig. 2b**. For comparison with actual CLIA signals, optical power from 100 pg mL^−1^ aldosterone (blue line) was measured (**Fig. 2b, Suppl. Fig. 3, Methods**).

**Figure 2:**
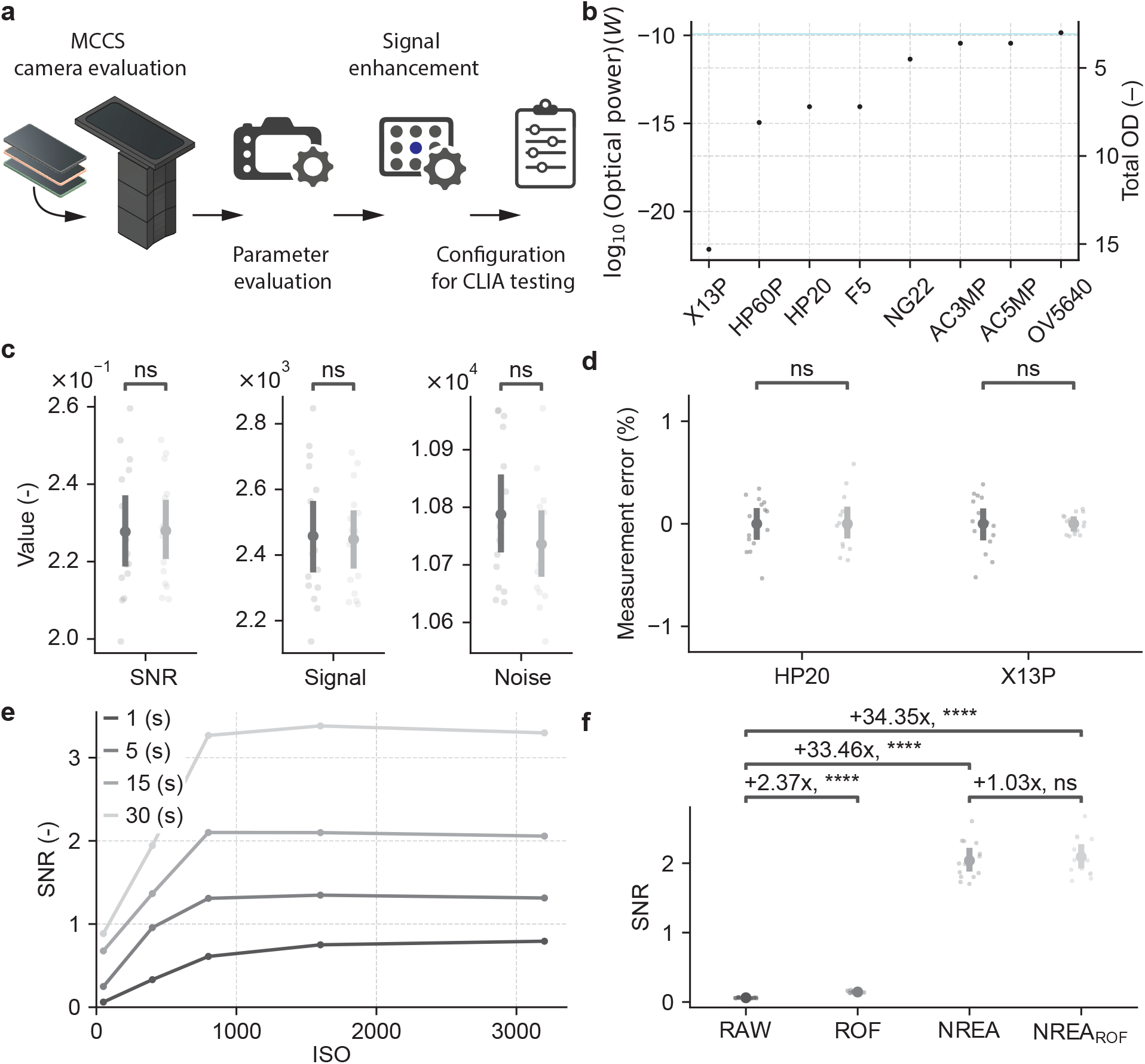
Camera Sensor Evaluation with MCCS. **a** Workflow using the MCCS to test configurations for CLIA testing. **b** Optical power limit for different cameras measured with a blue LED and varying numbers of neutral density filters (OD). Xiaomi 13 Pro (X13P), Huawei P60 Pro (HP60P), Huawei P20 (HP20), Fairphone 5 (F5), Nokia G22 (NG22), Arducam 3MP (AC3MP), Arducam 5MP (AC5MP), and OmniVision 5640 (OV5640). For reference, the cyan line is the power output of the signal from an aldosterone CLIA (sample conc. 100 pg mL^−1^). **c** Comparison of our (dark grey) and the smartphone’s native (light grey) app’s signal-to-noise ratio (SNR), signal, and noise (*n* = 15 images). **d** Precision of signal quantification for signal (dark grey) and background (light grey) for the smartphones HP20 and X13P as the relative error (% of full-scale). **e** SNR as a function of image acquisition parameters using the X13P (ISO and exposure time (s)). **f** The effect of image processing algorithms on the SNR using the X13P with 17 OD 0.9 filters added above the LED, calculated for the raw signal (RAW, *n* = 225 average over 15 images for 15 sets), the ROF denoised signal (ROF, *n* = 225 average over 15 images for 15 sets), the NREA processed signal on RAW and ROF denoised images NREA_ROF_ (*n* = 15, each).

To ensure reproducible acquisition control and fair comparison, we used our custom app (**Fig. 1d, Suppl. Fig. 1**). Comparing X13P RAW output to its native app at ISO 3200 and 30 s exposure via signal-to-noise ratio (SNR) and MCCS (**Fig. 2c**), Welch’s t-tests (*n* = 15 images/group) yielded nonsignificant differences in SNR (*p* = 0.963), signal (*p* = 0.885), and noise (*p* = 0.231) with negligible effect sizes (Cohen’s d |0.017||0.448|), supporting equivalence. Consequently, we proceeded with our custom app, which provides complete sensor control for our experiments. Measurement error must be small versus intrinsic CLIA signal variability for accurate quantification. We then used the MCCS to compare the most sensitive sensor available (X13P) with an average sensor suitable for CLIA (HP20). We compared the relative errors (% of full-scale) between regions with signal (ROI), and those with background (BG) for HP20 and X13P sensors with 8 and 17 0.9 ND filters, respectively (**Methods, Fig. 2d**, *n* = 15). Both models, modern 1-inch and 7 years older 1/2.3-inch sensors, showed normal distributions (Shapiro-Wilk *p >* 0.05). Welch’s t-tests found no mean differences (both *p* = 1.00) with zero effect sizes, corroborated by the Mann-Whitney U test (*p* ≥ 0.80). Percent coefficients of variation (%CV) were low (< 2.31 %) overall, indicating low intrinsic system variability and the suitability of both models for reliable light quantification **Table 1**. Next, the impact of ISO and exposure time on measured SNR was assessed with 6 · 0.9 ND filters, LED’s blue channel at level 10 (6.6 *±* 0.02 *×* 10^−13^ W) for X13P and HP20 (**Fig. 2e, Suppl. Fig. 4a**). The signal increased rapidly with ISO up to 800, plateauing thereafter, whereas with exposure it rose monotonically (**Suppl. Fig. 4b**,**c**). Noise trends varied; X13P noise peaked at 5 s exposure and ISO 400 then dropped with exposure at 30 s and ISO from 800 (**Suppl. Fig. 4d**); HP20 noise slightly rose with ISO up to 400 or 5 s exposure but remained stable otherwise (**Suppl. Fig. 4e**). Overall, SNR trends were comparable between phones.

To demonstrate how the MCCS can be used to evaluate novel image processing algorithms, we assessed the effect of Rudin-Osher-Fatemi (ROF) denoising and Noise Reduction through Ensemble Averaging (NREA) on SNR^44,45^. For images of a simulated weak CLIA signal (17 · 0.9 ND filters, LED’s blue channel at level 10) just above BG taken on X13P, the combination of the two, NREA_ROF_ improved SNR 34.35-fold while ROF or NREA alone increased it 2.37- and 33.46-fold (**Fig. 2f, Table 2**). Statistical tests confirmed significant enhancements (*p <* 10^−14^). NREA_ROF_ to NREA showed a 1.03-fold SNR gain (*p* = 0.579) with a rate change after 6 images (**Fig. 2f, Suppl. Fig. 5**). Overall, our algorithm NREA_ROF_ showed maximal SNR gain, and we used it going forward for maximal sensitivity.

Finally, testing if our approach boosts HP20 signals to X13P levels using the renin assay (**Suppl. Fig. 6a-g**), NREA and NREA_ROF_ raised HP20 SNR by 5.39–13.36 and 5.18–12.54-fold vs X13P RAW. Using the MCCS and only two images, the older sensor (HP20) outperformed X13P RAW 9.45-fold with NREA_ROF_, demonstrating the potential for signal processing to enhance weak signals captured by older, cheaper, and more readily available cameras.

### 2.3 Translation of the *in silico* MCCS results to CLIAs quantified using the MR

Our next aim was to demonstrate the utility of the method using a real-world example. For this, two CLIAs to determine the Aldosterone Renin Ratio (ARR) were chosen. ARR is a biomarker used in the diagnosis of Primary Aldosteronism (PA)^38^. To this end, we developed two magnetic microparticle-based CLIAs (**Fig. 3a**), one for the 37 kDa protein renin^46^ and the other for the 360 Da mineralocorticoid steroid aldosterone. These were chosen; 1) because the two assays feature two distinct immunoassay formats: a sandwich ELISA format for renin and, due to its small size, a competitive ELISA format for aldosterone and, 2) because the low abundance of renin in human blood (average plasma renin activity (PRA) for healthy donor samples 1.4 (0.2 - 5) ng mL^−1^ h^−1^ which equals 10.64 (1.52 - 38) pg mL^−1^ using a conversion factor of 7.6^47^, Patients with PA 0.2 (0.2 - 0.7) ng mL^−1^ h^−1^, 1.52 (1.52 - 5.32) pg mL^−1^ and patients with hypertension 1.6 (0.2 - 50) ng mL^−1^ h^−1^, 12.16 (1.52 - 380) pg mL^−1^ ^48^) requires a highly sensitive assay to be clinically relevant. We streamlined both assay workflows to achieve a short time-to-result of 35 minutes and to use standard laboratory consumables. To do this, a cost-effective 3D-printed, magnetic microparticle transfer tool was developed (**Fig. 3b, Fig. Suppl. Fig. 7, Methods**).

**Figure 3:**
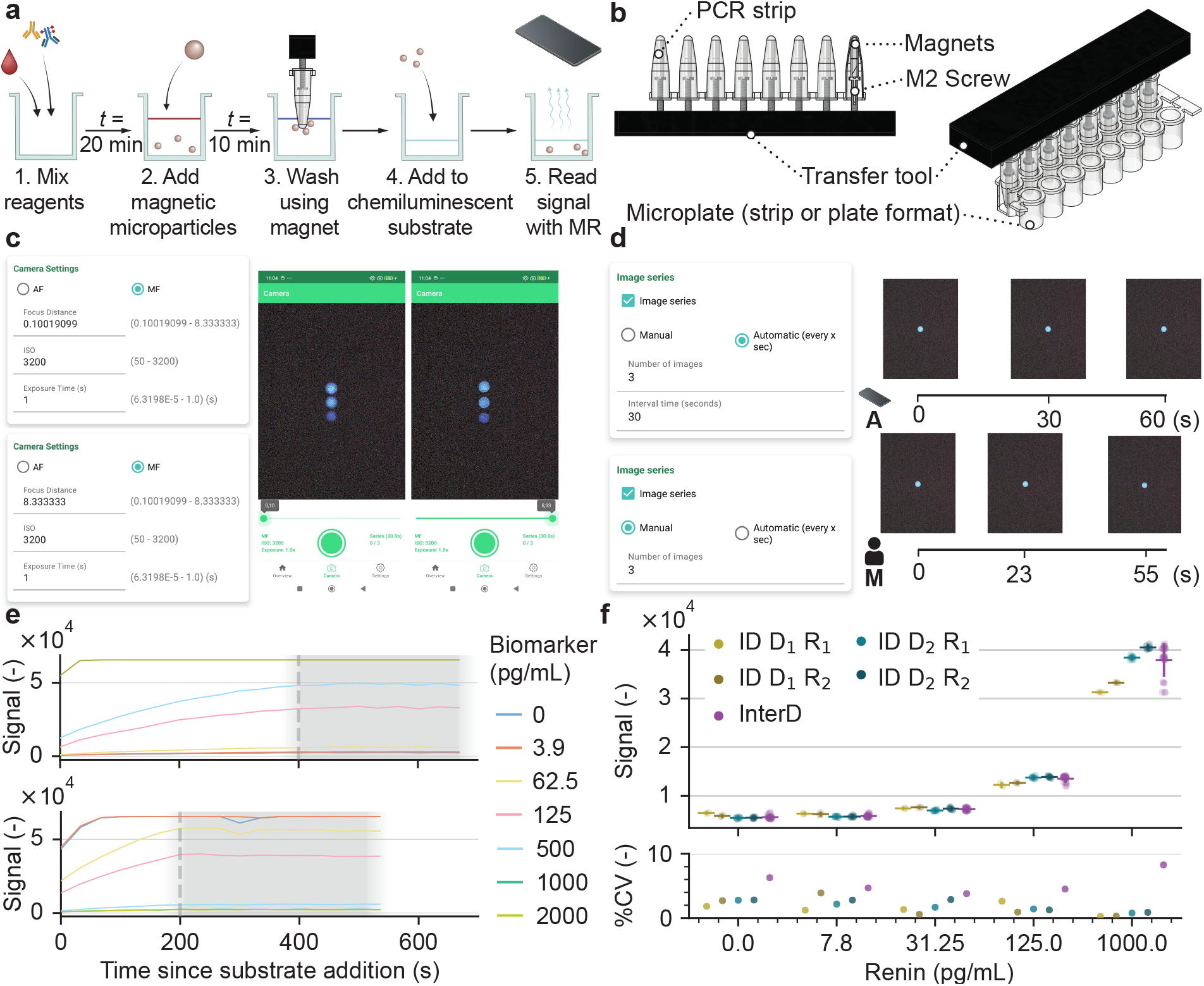
A method for rapid CLIA development and characterization using the MR device. **a** Schematic of the key steps and timings for the magnetic microparticle-based CLIA used in these studies. **b** 3D-rendering of custom-built magnetic microparticle transfer tool compatible with ANSI/SLAS Microplate Standard 96-well microplates or equivalent 8-well strips. **c** Focus control of our app, demonstrating out-of-focus and in-focus images using the MCCS. **d** Time-lapse capabilities of our app with fixed intervals in automatic mode (Automatic) or flexible interval defined by user (Manual) and resulting images acquired with MCCS. **e** Time dependence of signals for a renin (top) and aldosterone (bottom) standard curve with the start time of the signal reaching steady state (dashed line, on average 200 and 400 s for aldosterone and renin, respectively) and period during which the signal remains stable (grey box). **f** Intra-day (ID) and inter-day (InterD) measured on different days (D) with two technical replicates (R), assay signal and coefficient of variation (%CV) measured with the MR for five concentrations of a renin standard curve.

To reliably detect subtle CLIA signal variations, reproducible focus is crucial. We added focus control to fix autofocus distance and ensure consistent light collection from the same image plane across experiments (**Fig. 3c**). Another challenge is determining when the assay reaches maximum signal and its stable duration during assay development. To address this, we developed a time-lapse function in our app that enables automatic imaging at arbitrary intervals and durations across different smartphones (**Fig. 3d**). For our renin and aldosterone CLIAs, we measured the signal over time (**Fig. 3e**). After substrate addition, we found that the renin and aldosterone assays reach their signal maximum around 400 s and 200 s, respectively, and remain stable for at least 250 s (**Suppl. Fig. 8, Suppl. Fig. 9**). With this information in hand, we set out to determine the reliability of the CLIA signal quantification within our MR platform across multiple image acquisitions of assays, measured during the stable signal period. Tested on two different days (D_1_, D_2_), we used the renin assay with four concentrations and a blank (blank, 7.8, 31.25, 125, and 1000 pg mL^−1^) and two technical replicates (R_1_, R_2_) (**Fig. 3f**). We found, on average, a %CV = 1.77 *±* 1.0 for the MR intra-day (ID) variability (*n* = 3 and 11), indicating that the MR performed as expected and similarly with the assays compared to the MCCS *in silico* with a signal just above background (**Fig. 2c**). We observed larger variability in inter-day (InterD) and replicate comparisons with %CV = 5.51 *±* 1.8 (*n* = 28), indicating higher assay-dependent variability than the precision of the MR in quantifying a light signal (**Fig. 3f, Fig. 2c**).

### 2.4 Image processing for increased assay sensitivity and readout

To validate acquisition and pre-processing parameters identified *in silico* with MCCS, we tested them on our renin CLIA (**Fig. 4**). The luminol peak emission at 450 nm, with a central wavelength of 480 nm, overlaps well with the average blue channel sensitivity of mobile image sensors^49^ (**Fig. 4a**). Although 30 s exposure and ISO 3200 gave the highest *in silico* SNR under low light, brighter signals risk sensor saturation. We aimed to find parameters that balance low-light sensitivity and avoid sensor saturation across the clinically relevant range. Dynamic range was defined as between the lower limit of detection (LLoD) and the upper limit of detection (ULoD) for renin and aldosterone assays (grey box, **Fig. 4b-e**). The green channel showed higher dynamic range but an elevated LoQ (**Fig. 4b**), and at ISO 1600, it best discriminates the signal from low concentration samples (**Fig. 4c**). At ISO 1600, 30 s exposure, all measured concentrations were above the LLoD; whereas for 5 s, 7.8 pg mL^−1^ was outside the LloD (**Fig. 4d**). Motivated by the good performance of our signal processing on the SNR (**Fig. 2f**), we asked if the LLoD for ch = green, ISO 1600, t_exp_ = 5 s can be improved and reach the performance of the 30 s single exposure. We took six serial images with those settings and applied NREA processing. Single images with NREA_RAW_, and NREA_ROF_, improved the lowest concentration above LoQ, unlike RAW or ROF alone (**Fig. 4e**). Six images cumulatively increased the dynamic range and detectability of low concentrations, outperforming a single 30 s exposure, even for aldosterone, which exhibits higher variability (**Fig. 4d,e, Fig. Suppl. Fig. 10a-d**).

**Figure 4:**
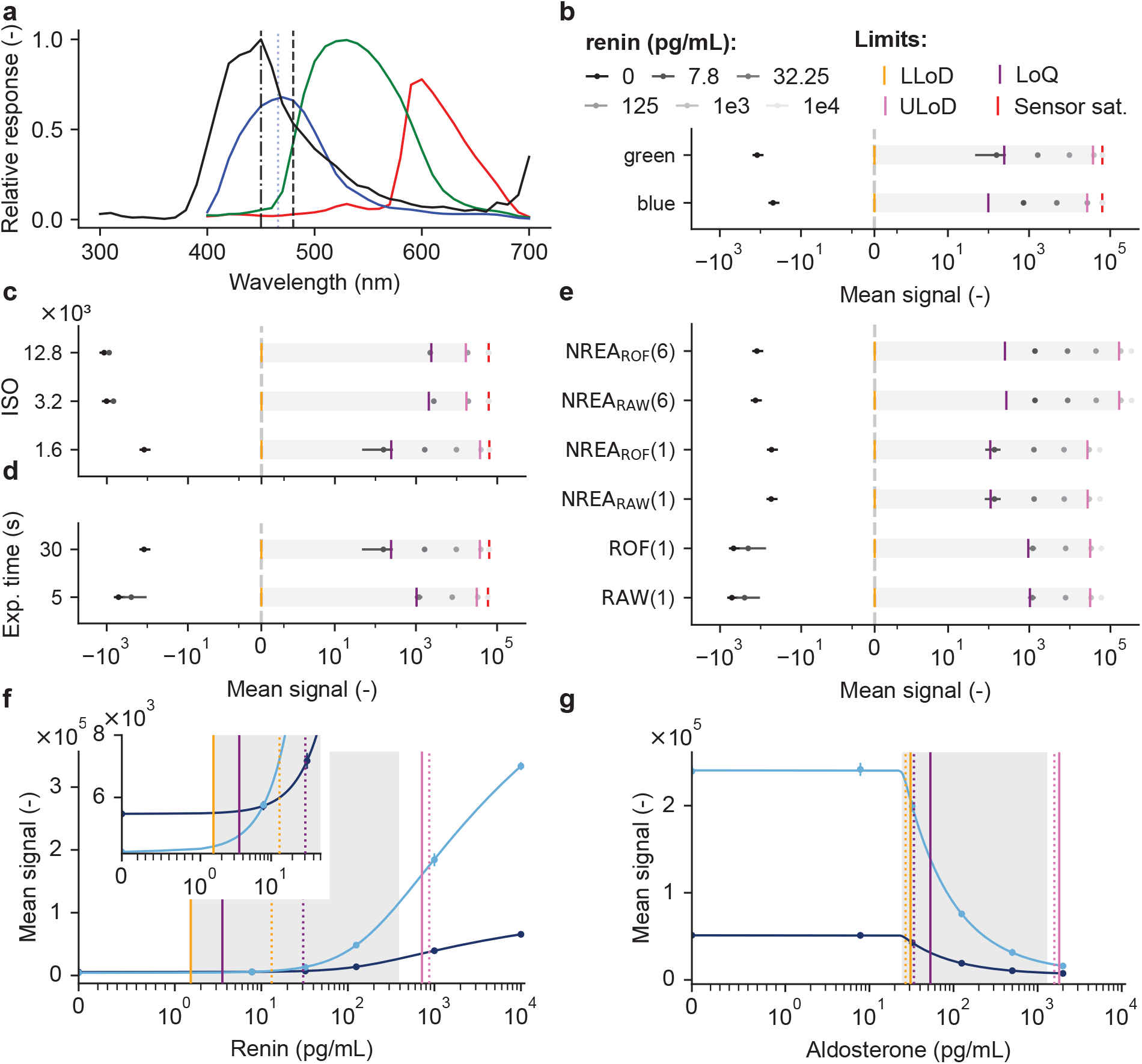
The effect of image acquisition and processing on the lower limit of detection (LLoD), limit of quantification (LoQ), and upper limit of detection (ULoD) on two CLIAs quantified with MR and X13P. **a** Average spectral response of smartphone camera sensors in blue (line), green (line), and red (line) channels (data taken form^49^) with Luminol emission spectrum (black line) and its peak and center wavelength (vertical dotted dashed line and dashed line). **b** Primary color channel-based dynamic range for a renin assay at different concentrations with the average signal (black dot, vertical line (mean *±* SD), *n* = 2) normalized to the LLoD (vertical orange line) with LoQ (vertical violet line), ULoD (vertical pink line) and sensor saturation (vertical dashed red line, at 16-bit). The grey box indicates the range between LLoD and ULoD. The further apart the data and limits, the larger the dynamic range. **c** Same as in **b** with the green channel and different ISO values (1600, 3200, 12800). **d** Same as in **b** with the green channel, ISO 1600 for different exposure times (t_exp_ = 5, 15, 30 s). **e** Same as in **b** with the green channel, ISO = 1600, t_exp_ = 5 s for unprocessed (RAW), ROF denoising (ROF), NREA on RAW or ROF denoised images (NREA_RAW_, NREA_ROF_) for one and a series of six images. **f** Standard curve for renin for RAW (dark blue dots, vertical bar, mean *±* SD), and NREA_ROF_ (light blue dots, vertical bar, mean *±* SD) with LLoD, LoQ, and ULoD (vertical orange, violet, and pink line, dotted RAW, solid NREA_ROF_). The inset shows the zoomed-in region between 0 and 50 pg mL^−1^, visualizing the improvement in sensitivity of NREA_ROF_ between 0 and 10 pg mL^−1^ compared to the other condition. **g** Same as in **f** for the standard curve of the aldosterone assay.

Standard curves for renin and aldosterone with RAW (dark blue) and NREA_ROF_ (light blue) confirmed our findings (**Fig. 4f,g**). To determine the LLoD, LoQ, and ULoD, we followed FDA/ICH guidelines^41,42^ and fit the simplest model to the standard curves (linear, 4PL, and 5PL, **Methods**). The 5PL model best fit all data and confirmed LLoD, LoQ, and ULoD, **Table 12**. For renin, with relevant concentrations as low as 1.52 pg mL^−1^, our NREA_ROF_ signal processing covered the entire relevant range with LloQ at 1.53 pg mL^−1^, while RAW is approx. a factor of nine higher (**Table 8**). This surprisingly shows our mobile reader (MR), built with off-the-shelf parts and a smartphone, rivals bulky, expensive SOTA readers (renin PR LLoD 4.01) (**Suppl. Fig. 11a, Table 8**). _CLIA_MDK also showed that the aldosterone assay could not fully cover its range due to variability in blank signals (**Fig. 4g**), with MR and PR again performing comparably(**Suppl. Fig. 11b**).

### 2.5 Pilot study on human samples correlating MR with reference method

To further evaluate the performance of our platform (MR), we utilized characterized patient samples obtained from a commercial provider (Labcorp) in a pilot study with our example assays. We measured standard curves for each renin and aldosterone using spiked plasma (renin / prorenin double-depleted plasma) and human serum, respectively. We chose six concentrations, including a blank, covering the clinically relevant range^47,48^ for both markers (**Fig. 5**).

**Figure 5:**
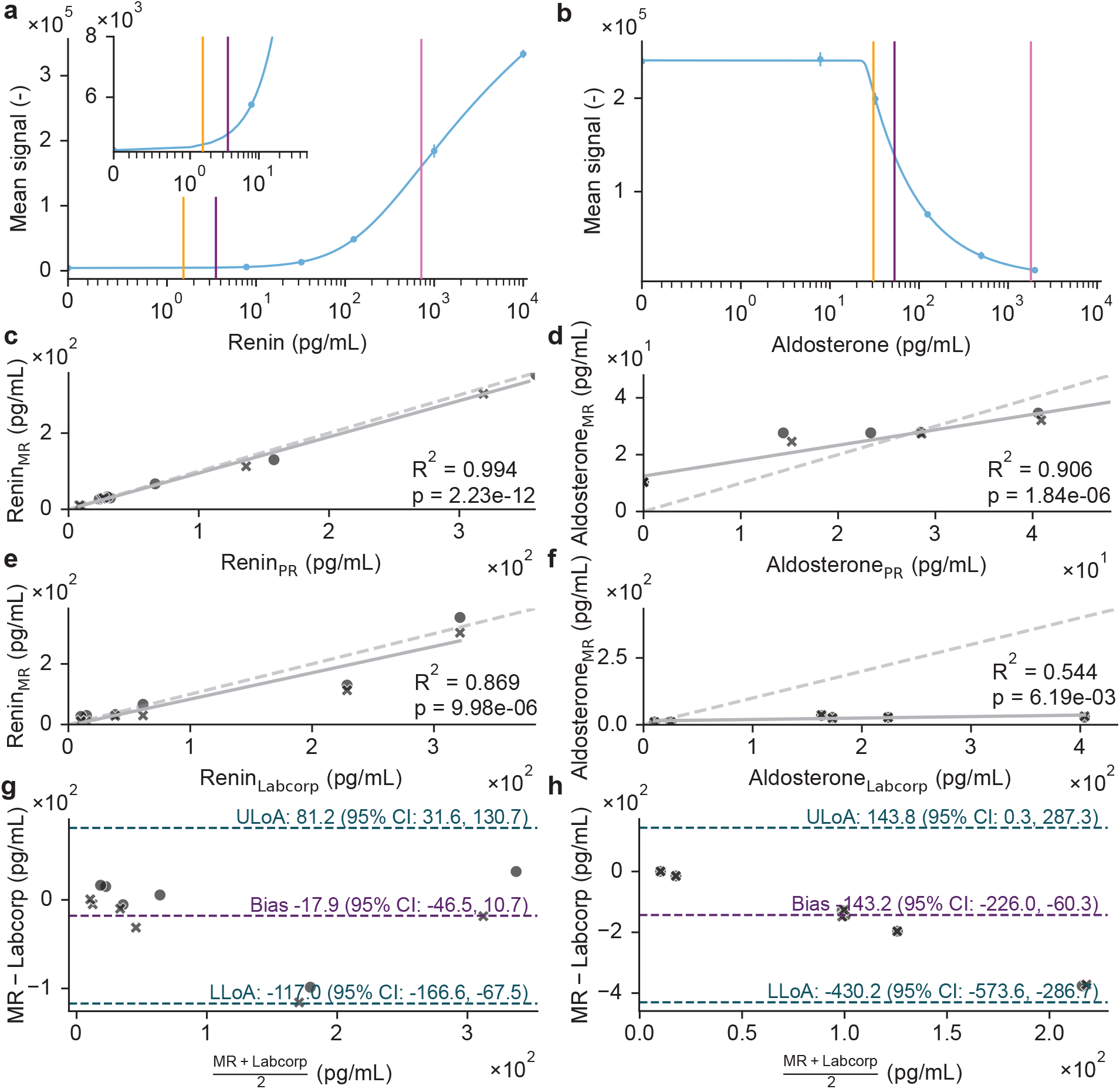
CLIAs measured with MR in comparison with SOTA (PR, and LC/MS Labcorp) using patient samples. **a** Renin standard curve with a five-parameter logistic (5PL) fit (blue line) with LLoD, LoQ, and ULoD (vertical orange, violet, and pink lines, respectively) on proposed signal processing. The standard curve was measured using spiked renin/prorenin double-depleted plasma, measured with two technical replicates. **b** same as in **a** but measured in aldosterone spiked human serum. **c, d** MR correlation with PR measurements and Pearson correlation fit (solid line) for renin and aldosterone, respectively (*n* = 6) measured in two technical replicates (crosses and dots), identity (dashed line). **e, f** same analysis as in **c, d** correlating MR using independent LC/MS-based methods provided by Labcorp. **g, h** Bland-Altman plot comparing the MR results with Labcorp results for renin and aldosterone, respectively.

We fitted a 5-parameter logistic regression (5PL) model and found that for both standard curves, the 5PL fit explained a large portion of the data’s variance (renin, *R*^2^ = 0.99; aldosterone, *R*^2^ = 0.90) (**Fig. 5a,b**). We measured a small set of patient samples (*n* = 6) using the MR and applied the same processing parameters as those used for the standard curves. Renin was quantified in plasma; aldosterone in serum in a paired set of samples. The quantified signals from patient data were then converted to concentrations using the standard curve 5PL model. They were found to be in good agreement with the results of the SOTA PR, for renin (*R*^2^ = 0.994, *p* = 2.23*e* − 12) and aldosterone (*R*^2^ = 0.906, *p* = 1.84 − 6) (**Fig. 5c,d**). We found strong agreement between the MR and the SOTA measurement technology from Labcorp (*R*^2^ = 0.869, *p* = 9.98*e* − 6) with a bias of -17.3 (95% CI -45.6, 10.7) pg mL^−1^ (**Fig. 5e,g**). The PR was comparable but with a higher bias (*R*^2^ = 0.913, *p* = 1.27*e* − 6, bias of -13.0 (95% CI -36.4, 10.4) pg mL^−1^ **Suppl. Fig. 11c**,**e**. For aldosterone, however, we found poor agreement between MR and LC/MS methods (*R*^2^ = 0.544, *p* = 6.19*e*− 3) with a bias of -143.2 (95% CI -226.0, -60.3) pg mL^−1^ (**Fig. 5f,h**). Again, the PR yielded a comparable result with (*R*^2^ = 0.514, *p* = 8.73*e* 3) and a bias of -146.5 (95% CI -226.3, -66.7) pg mL^−1^ (**Suppl. Fig. 11d**,**f**). The Passing-Bablok (PB) regression revealed no significant proportional bias between the MR and PR for renin; however, a slight constant bias, slope, and intercept, 0.94 (95% CI=[0.80, 0.99]) and 4.47 (95% CI=[0.55 to 7.99]) pg mL^−1^. For aldosterone, there is a proportional and constant bias, with slope, and intercept, 0.54 (95% CI=[0.34, 0.62]) pg mL^−1^ and 11.12 (95% CI=[10.34 to 18.16]) pg mL^−1^ (**Suppl. Fig. 12a**,**b**). This demonstrates an assay dependent near-equivalent performance between our approx. 20x cheaper MR compared to the commercially available PR.

The same analysis, compared to Labcorp, reveals broader CI, suggesting that the estimate is imprecise, possibly due to limited or variable data, but no proportional and constant bias for renin (**Suppl. Fig. 12c**,**e**). In contrast, PB regression for aldosterone yielded slopes of 0.05 (95% CI=[0.0 to 0.09] pg mL^−1^ and 0.10 (95% CI= [0.0 to 0.13] pg mL^−1^, with intercepts of 13.88 (95% CI=[8.80 to 27.51] pg mL^−1^ and -0.97 (95% CI= [–3.34 to 19.04] pg mL^−1^ indicating significant proportional bias between MR and PR to Labcorp and small constant bias (**Suppl. Fig. 12d**,**f**). The poor correlation between the MR and PR CLIA and the LC/MS method from Labcorp is known due to different measurement principles^50–52^. Given the small sample size of *n* = 6 patient samples, further performance evaluation is necessary.

### 2.6 Performance validation of MR against reference methods in a larger cohort

Although our pilot study of human samples (*n* = 6) showed high correlation between MR and both reference methods (PR, *r* = 0.999, Labcorp, *r* = 0.952, **Table 9**) for the renin assay, the limited sample size precluded robust estimation of true assay performance. Power calculations indicated that at least 50 samples are required to estimate the actual correlation with both statistical significance and clinical relevance (**Methods**). Accordingly, we undertook an analytical performance study with an expanded cohort to assess both renin and aldosterone assays with the MR. We interpret the data primarily as a localization experiment: MR–PR agreement tests reader equivalence, while comparison to LC–MS serves as an orthogonal method comparison that can reveal assay- and matrix-dependent limitations.

We first acquired standard curves with six replicates for twelve spiked sample concentrations, including a blank for renin and aldosterone, respectively (**Fig. 6a, Suppl. Fig. 13a**). Compared to the pilot study, we observed a higher LLoD due to differences in blank signals (**Fig. 5a,b, Table 8**). Next, we measured 50 human patient samples with three technical replicates (same reagent mixes, same day) for both assays. For renin, excellent correlation between the MR and PR (*R*^2^ = 0.990, *p* = 3.57*e* − 151) was confirmed. Even stronger correlation was observed with the SOTA reference method (Labcorp) for both MR (*R*^2^ = 0.934, *p* = 3.25*e* − 89) and PR (*R*^2^ = 0.931, *p* = 1.04*e* − 87), with narrower confidence intervals than in the pilot study (**Fig. 6c,d, Table 9**). This demonstrates that our assay performs with robust precision and minimal uncertainty across the clinically relevant range, comparable to both PR and the reference standard. Bland-Altman analysis comparing MR to PR revealed a small bias of 24.0 (95% CI=[-16.7, 31.2]) pg mL^−1^ with overestimation occurring with high concentration samples (≥ 600 pg mL^−1^) (**Fig. 6d, Fig. Suppl. Fig. 14a**), confirming the overall interchangeability of MR and PR (**Fig. Suppl. Fig. 14b-d**) in the clinically relevant ranges^47,48^.

**Figure 6:**
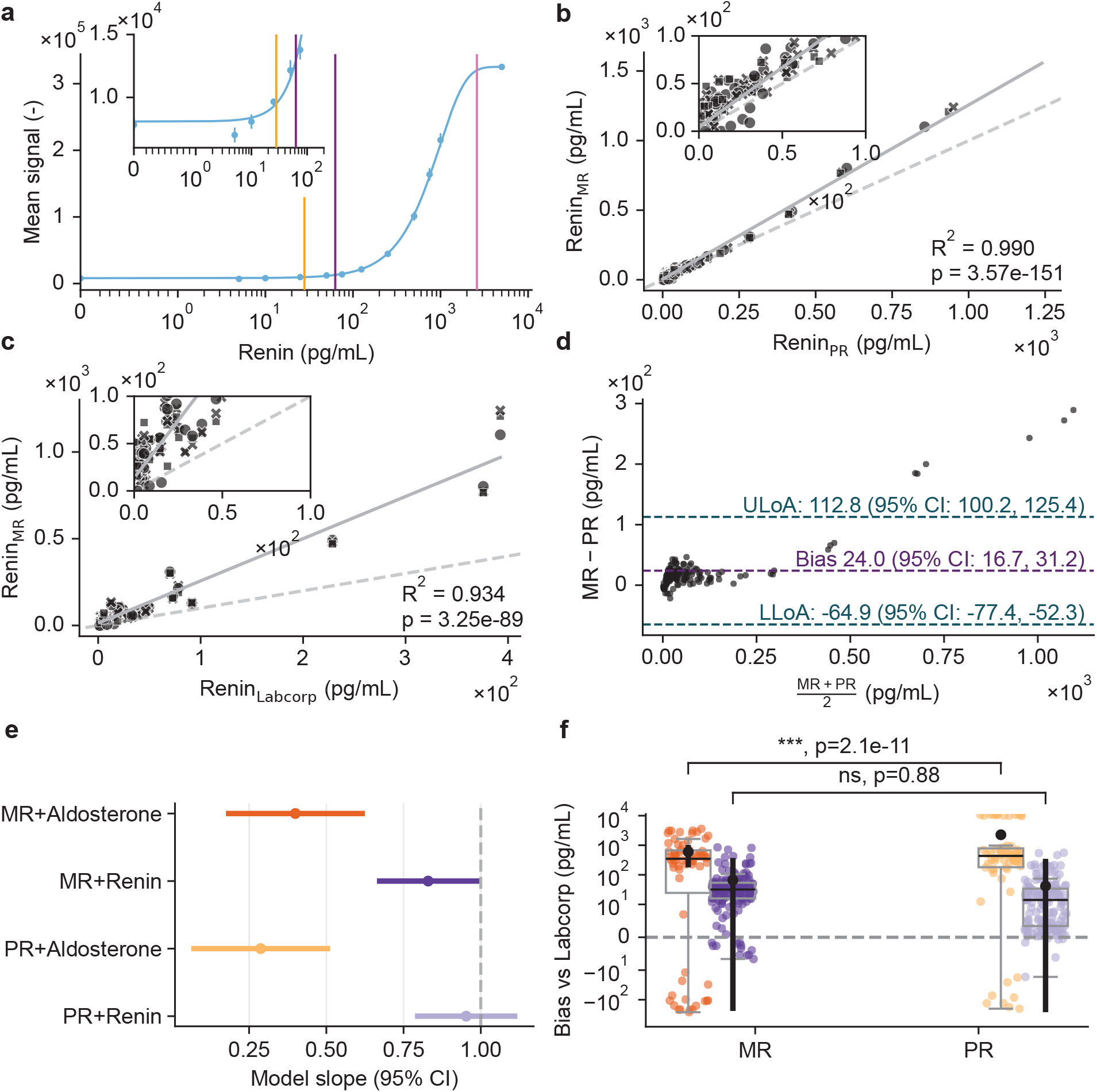
Performance study comparing MR with SOTA using patient plasma samples on the renin assay. **a** Renin standard curve with 5-parameter logistic regression (5PL) fit (blue line) with LLoD, LoQ, and ULoD (vertical orange, violet, and pink lines) recorded with MR with proposed signal processing (n=6 per concentration). The standard curve was measured using spiked renin/prorenin double-depleted plasma. Vertical lines show LoD (yellow), LLoQ (purple), ULoQ (pink). **b** Correlation of MR to PR measurements of patient plasma-EDTA samples (*n* = 50), measured in technical triplicates (circle, square, and cross shapes), linear fit (grey line) and identity line (grey dashed line). **c** Same as in **b** for the correlation of MR renin quantification with an independent method (LC/MS, Labcorp) for plasma samples (*n* = 50). **d** Bland-Altman plot comparing the MR results with PR measurements (*n* = 50). **e** Forrest plots showing slope estimates of a mixed effect model (see **Methods**) with horizontal error bars, estimated slopes (calibration model) with mean (dot) and 95% CI (line) for each device × assay combination (MR+Aldosterone (orange), MR+Renin (violet), PR+Aldosterone (light orange) and PR+Renin (light violet)). Slopes near 1 indicate proportional agreement (grey dashed line) which supports equivalence for the Renin assay between MR and PR. **f** Actual data (scatter, colors as in **e**) and boxplots of measured minus reference concentrations by device × assay with overlaid model-based estimates (mean, black dot) and 95% CI (black vertical line). The box represents the inter-quartile range (IQR), with the median shown as a horizontal black line. Whiskers extend to the most extreme data points within 1.5 times the IQR. Asterisks indicate significance of bias relative to zero (grey dashed line); (*n* = 432)

To disentangle device- and assay-specific effects from biological sample variability, we applied linear mixed-effects models using the Labcorp measurements as an independent reference (**Fig. 6e,f, Fig. Suppl. Fig. 16a-c, Table 10**). Calibration analysis revealed assay-dependent proportional bias: aldosterone slopes deviated from unity on both devices, whereas renin slopes were close to 1 with overlapping confidence intervals, supporting proportional agreement and device interchangeability (**Fig. 6e**). Absolute bias analysis confirmed that aldosterone measurements exhibited significant positive bias on both devices, with larger bias on PR than MR (PR–MR: *p* = 2.1 *×* 10^−11^), while renin measurements showed no significant bias relative to the reference and no device-related differences (PR–MR: *p* = 0.88) (**Fig. 6f**). For the competitive assay targeting aldosterone, the MR and PR remained strongly correlated (*R*^2^ = 0.812, *p* = 6.27*e* − 25) (**Fig. 5d, Suppl. Fig. 13b**), but both showed poor correlation (MR (*R*^2^ = 0.056, *p* = 0.0564) and PR (*R*^2^ = 0.036, *p* = 0.126)) to the orthogonal LC/MS method (Labcorp) due to intrinsic differences between immunoassay and LC/MS reference method (**Suppl. Fig. 13c**,**d**). MR and PR perform similarly on the assay (**Fig. Suppl. Fig. 14e**,**f, Suppl. Fig. 13e**,**f**) and Bland-Altman analysis revealed a bias of 73.0 (95% CI=[-161.1, 15.1]) pg mL^−1^ (**Fig. Suppl. Fig. 14f**). The PB regression yielded a small but significant proportional and constant bias for renin between MR and PR (slope=1.08, 95% CI=[1.03, 1.13]; intercept=12.38, 95% CI=[10.16, 13.90]) (**Suppl. Fig. 15a**), whereas aldosterone showed slope=0.83 (95% CI=[0.62, 1.03]) and intercept=7.59 (95% CI=[-107, 110]), indicating method equivalence (**Suppl. Fig. 15b**). MR versus the reference method (Labcorp) confirmed pronounced bias for renin (PB slope=2.07 95% CI=[1.95, 2.30]; intercept=16.02 95% CI=[13.28, 19.75]) (**Suppl. Fig. 15c**) and for aldosterone (slope=0.84 95% CI=[0.17, 1.37]; intercept=406.92 95% CI=[270, 520]) (**Suppl. Fig. 15d**). PB for PR versus Labcorp showed slightly worse performance than than MR (**Suppl. Fig. 15e**,**f**). Across both aldosterone and renin, the ICC for sample-level variance was 0.21, indicating that one-fifth of total variance arises from biological differences between samples, with the remaining 79% attributable to measurement-level variability (**Table 10**). Overall, these results confirm that device differences are minor relative to biological variation, supporting the interchangeability of MR and PR.

## 3 Discussion

We demonstrate a modular platform for rapid and systematic evaluation of mobile camera sensors in quantitative chemiluminescent immunoassay (CLIA) applications and CLIA assay development. Importantly, the platform _CLIA_MDK, built from affordable, commercially available components, delivered analytical performance equivalent to a state-of-the-art plate reader (PR, 20-50k USD), but at a dramatically lower cost (< 40 USD materials plus smartphone (100-1000 USD)). Key advances include *in silico* sensor and parameter screening using the MCCS, followed by laboratory validation of the candidate sensor configuration. Together, the renin (sandwich) and aldosterone (competitive) assays serve as complementary stress tests of the platform, enabling _CLIA_MDK to localize performance bottlenecks to either the reader (sensor/acquisition/processing) or to assay- and matrix-dependent effects. Beyond a single assay, _CLIA_MDK provides a generalizable end-to-end workflow linking *in silico* sensor screening to standardized raw acquisition and validated quantitative readout. This accelerates the selection of a suitable sensor and its configuration, reducing the use of precious samples and expensive reagents in the development of novel assays. The system’s modular hardware can be locally produced, easily adapted, facilitating future extensions (e.g., microfluidic base modules) and resource optimization. Notably, image-based quantification not only supports multiplexed detection^53^ but also optimizes sensitivity by leveraging spectral differentiation.

*In silico* evaluation of five smartphone cameras and three low-cost CSI cameras showed all smartphone models outperforming the CSI sensors in sensitivity. Our custom app enables standardized raw data acquisition and comparison across devices, validating consistent signal and noise handling. Both older (HP20) and newer (X13P) smartphone sensors performed reliably, with low measurement error and high suitability for CLIA quantification as per FDA guidelines^41^. Our platform further automates the essential step of signal processing algorithm evaluation, especially for targeting low-abundance biomarkers with weak signals. Combining ROF denoising with NREA significantly improved SNR for signals near the noise level, consistent with IUPAC and imaging guidelines for minimal SNR in reliable quantification^54–56^. While post-processing cannot change a sensor’s photon-to-digital conversion capability, we showed advanced processing can render older sensors suitable for CLIA detection. Notably, the HP20 smartphone (2018) achieved a 9.3-fold SNR gain over raw output from a contemporary 1-inch sensor using only two images. This highlights the impact of post-acquisition processing and demonstrates that even older, widely available smartphone cameras can serve as viable platforms within modular hardware designs.

To illustrate _CLIA_MDK’s application, we developed two example magnetic microparticle-based CLIAs targeting ARR^57^, as rapid tests are lacking for these difficult-to-detect small molecules. Magnetic microparticles were selected to immobilize capture reagents for their favorable mass-transport and surface properties, enhancing assay performance and shortening incubation times^58^. With our cost-effective microparticle handling tool, we simplify test handling during development. Given the intrinsic time-dependent signal variation of CLIAs, our mobile app introduces time-lapse monitoring to identify steady-state signal plateaus and improved focus control for reproducible acquisition, accounting for sensor repositioning between experiments. The system’s %CV from sensor, app, or processing was lower *in silico* than inter-day assay variability *in vitro*, highlighting the platform’s reliability and importance of optimizing acquisition parameters per analyte.

Spectral analysis confirmed that luminol emission aligns with the sensor’s blue channel; additionally, multi-channel acquisition and primary color channel analysis expanded the quantifiable dynamic range. This, combined with optimal acquisition parameters and signal processing, effectively elevated the lowest measurable concentrations above the LoQ. Most importantly, the MR achieved LLoDs similar to those of the SOTA plate reader (PR), and variability in blank measurements identified nonspecific binding as a current bottleneck (**Table 8**). With the lowest achieved LLoD of 1.6 pg mL^−1^ for renin, our MR platform is highly sensitive. Our workflow rapidly evaluates the parameter space to confirm *in silico* translation to assays.

Pilot testing with patient samples further confirmed the good correlation of MR and SOTA PR for both assays. The renin assay showed robust agreement with certified laboratory analysis, with only minor bias. Lower performance in the aldosterone assay was consistent with literature-reported differences between competitive CLIA and LC/MS reference methods^59,60^. Critically, the pilot study validated the utility of *in silico* MCCS sensor selection and platform performance on clinical samples, justifying expansion to a larger cohort for statistical power and generalizability.

The performance study’s results, including 50 patient samples, further support the MR’s reliability: strong readout consistency, low SD, and high correlation with both SOTA methods, PR and Labcorp, were achieved for the renin assay and mixed-effects modeling confirmed that device differences were minor relative to biological sample variability, supporting interchangeability; importantly, excellent agreement between our CLIA (MR) and Labcorp’s reference method (*r* = 0.97, 95% CI: 0.95–0.98; *p* = 3.3*e* − 89), with a narrower CI than targeted, reflects increased statistical power and robustness, supporting the validity and potential clinical utility of the assay and ability of _CLIA_MDK to detect assay-specific issues early. Bland–Altman analysis of MR versus PR revealed slight bias from highly concentrated samples outside the clinically relevant range and biological variability rather than sensor or quantification limits. For aldosterone, higher replicate variability compared to the pilot study, likely reflects intrinsic assay and sample-specific confounding factors, like enzyme substrate depletion, mixing, or thermal effects – known immunoassay interference contributors^50,60–63^ – but both MR and PR were similarly affected, indicating confounders inherent to the competitive assay format rather than device limitations. Differences in bias relative to the independent reference laboratory method are expected as Labcorp’s renin activity assay relies on LC/MS detection of angiotensin I, while immunoassays target direct antigen measurement, generating systematic differences well documented in the literature^64^ and evidenced in the range in conversion factors (**Table 4**). Aldosterone quantification also varies between LC/MS and immunological methods^59,60^.

Successive smartphone generations have made more sensitive CMOS sensors broadly available. Our findings validate using these sensors in a modular platform, demonstrating mobile devices as a cost-effective alternative to conventional state-of-the-art plate readers or controllers of dedicated sensors. Using two new CLIA assays targeting key biomarkers of primary aldosteronism, the platform quantified analyte concentrations within the clinical range, achieving an LLoD below 1.6 pg mL^−1^ and comparable performance to reference methods for renin, while also identifying the need for further assay optimization or a different assay format for aldosterone^65^. As with any diagnostic approach, performance validation remains essential for each assay and sensor combination. Collectively, these results establish _CLIA_MDK as a high-quality and accessible tool to accelerate assay development and expand access to sensitive diagnostics. If widely implemented, _CLIA_MDK could help lower healthcare system costs while providing a scalable framework that expedites the evaluation of new signal-processing and integration of additional sampling and assay modalities, such as microfluidics. Beyond using the platform for low-cost CLIA development, we envision validating _CLIA_MDK as a foundation to extend laboratory-grade analyses into more diverse and globally accessible healthcare settings.

## 4 Materials & Methods

### Design and assembly of MCCS and MR

The modular camera calibration system (MCCS) and mobile reader (MR) were designed with Autodesk Inventor Professional (Autodesk, USA). RGB LEDs were used (WS2813, NeoPixel Buttons, Adafruit, USA). Diffusor plates were laser cut from Opaque Acrylic, 3 mm. Neutral density (ND) filters were laser cut with a diameter of 12.5 mm for lowering the LED intensity barely above the detection limit using 0.3ND (LEE Filters, Art. B209, UK) and 0.9ND (LEE Filters, Art. B211, UK). The diameter was chosen to be compatible with widely available filters and the 12.5 mm filter diameter standard. Two custom electronic boards were designed (see **Fig. Suppl. Fig. 17, Table 11**). An LED shield and the MCCS board to control the LEDs from a microcontroller over Bluetooth (nRF52840, Seeed Studio, CN). All the enclosure parts of the MR (MCCS and MR) were 3D-printed in-house using Onyx filament (micro carbon fiber-filled nylon). The internal structure was laser cut (2’’ or HPDFO, lens using standard settings, VLS2.30, Universal Laser Systems Inc., USA) from transparent PMMA sheets (2, 3, 4, 8 mm Plexiglass XT Technische Folien Farblos 99524, Plexiglass XT Allround Farblos 0A000, Röhm, Switzerland). The following painting system was applied to absorb stray light. Layer one was sprayed with primer (edding 5200 primer), layers two and three were sprayed black (edding 5200 black matt 4-5200901), and layer four was painted with 99.4% absorbing black (Musou black, MB-100, Koyo Orient Japan Co., Ltd., Japan). 3D-printed parts were also primed (Vallejo primer black VJ74602, Acryicos Vallejo, S.L., Spain) followed by 99.4% absorbing black (Musou black, MB-100, Koyo Orient Japan Co., Ltd., Japan).

### App development and camera sensor evaluation

The ChemiLuminescent ImmunoAssay Mobile Development Kit (_CLIA_MDK) includes our custom-developed mobile phone app written in Java for Android operating systems (Android Studio Ladybug, version 2024.2.1, Runtime version 21.0.3+-12282718-b509.11 amd64, VM: OpenJDK 64-Bit Server VM by JetBrains s.r.o., Gradle targetSDK 33). The MCCS control and the camera control are summarized in **Fig. Suppl. Fig. 1** and for the app (see **Data Availability**). The cameras of five smartphones were evaluated. Xiaomi 13 Pro (X13P), 221013G, MIUI 14.0.22.0 (TMBEUXM), Android version 13 TKQ1.220905.001, Huawei P20 (HP20), EML-L29/EML-L09, EMUI 10.0.0.190, Android version 10, 862934042305497, Huawei P60 Pro (HP60P), MNA-LX9, EMUI 13.1.0.183 (C432E5R2P3), Android version 12, 865607061858714, Fairphone 5 5G (FP5), FP5, 2a71cbce, Android version 13, FP5.TT3B.A.083. 20230728, Nokia G22 (NG22), A-1528, 00WW_1_240, Android version 12, AS0609H069P50601089. Furthermore, three CSI camera modules connected to an Xiao ESP32S3 Sense (Seeed Studio, CN) were evaluated and controlled through our app. A 3- and 5-megapixel SPI camera module, AC3MP and AC5MP (Arducam, CN), and the 5-megapixel CMOS image sensor OV5640 (OmniVision Technologies, Inc., USA).

### Image pre-processing and signal quantification

Custom scripts were written in Python (version 3.9.13) to preprocess the images using various denoising strategies. We evaluated the ROF denoising^44^ model and the NREA algorithm^45^ with custom Python implementations. The ROF model minimizes the total variation norm of the image while preserving fidelity to the original noisy image, reducing noise for both the X13P and HP20. The NREA method allows the signal to accumulate while suppressing noise. It first applies a circular averaging filter, acting as a low-pass filter to suppress high-frequency noise while preserving the LED’s circular signal. The image is then mean-subtracted to center the intensity distribution around zero, enabling accumulation of multiple transformed images to average out noise while retaining the signal. To then automatically detect the wells from images, we implemented custom algorithms to quantify the signal in each primary color channel (R, G, B) from the RAW image input. The circular wells are detected using a Circular Hough Transform^66^. Let the detected circle be represented by its center (*x*_*c*_, *y*_*c*_) and radius *r*. The equation of the circle is:

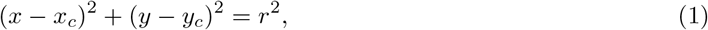

where (*x*_*c*_, *y*_*c*_) are the coordinates of the center, *r* the radius of the circle, and (*x, y*) the coordinates of a point on the circumference of the circle. From the center of the well (*x, y*), a square is defined with side length *s* satisfying *s*^2^ ≤ 2*R*^2^, ensuring it fits inside the circle. For all experiments, we used *s* = 60 pixels, unless stated otherwise. The top-left and bottom-right corners of the square are defined as: 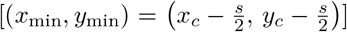 and 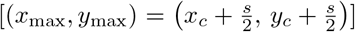. The set of pixel coordinates (*x, y*) that lie inside the square region of interest within the well *S* is given by:

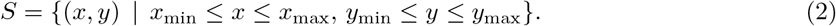

The background region *B* is defined as the set of pixel coordinates outside the circle:

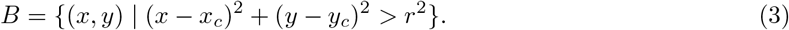

Let *I*(*x, y*) denote the pixel value at position (*x, y*) in the image *I*. The mean signal values in the square region *S* and the background region *B* are defined as

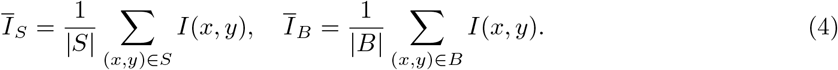

The raw signal is then defined as the mean signal value over the square region *S*,

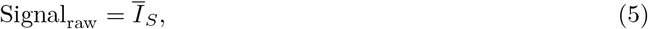

and the background-corrected signal as the difference between the mean signal in the well region *S* and the background region *B*:

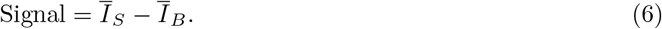

The noise is defined by combining the standard deviations of pixel signal values in *S* and *B*:

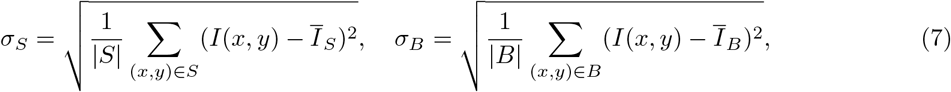

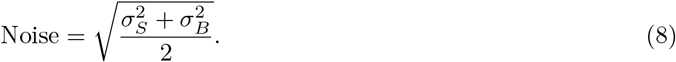

The background-corrected signal-to-noise ratio (SNR) is defined as:

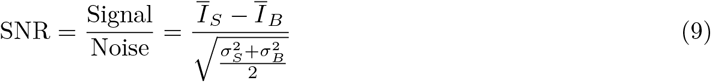

To measure the power of the LEDs we used a Thorlabs S130VC power meter (S/N 11040622), and Thorlabs PM100USB (Thorlabs Inc, USA).

### Determination of lowest detectable signal

The optical power of the LED signal (P_LED_) in the blue channel and its uncertainty was calculated by averaging the measurements (n=100) for when the LED is on and off according to *µ*_LED_ = *µ*_LED(10)_ − *µ*_LED(0)_ with 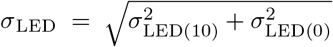. This resulted in a measured optical power P_LED(10)_ = 1.420 *±* 0.005 *×* 10^−7^ W at brightness 10 (a.u. PWM, on 8-bit input range (0-255)), with two diffusor plates (opaque white acrylic, 3 mm thickness) and without ND filters.

We used this LED optical power (P_LED(10)_) for all MCCS experiments in combination with neutral density (ND) filters (0.3ND (LEE Filters, Art. B209, UK); 0.9ND (LEE Filters, Art. B211, UK)) to incrementally reduce the incoming light to determine the lowest detectable signal of each camera sensor. Since the signal itself also fluctuates, we are interested in signal brightness at which *µ*_signal_ is as low as possible while still satisfying 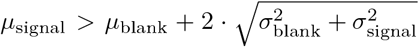 where *µ*_signal_ is the brightness of a blank measurement when the LED is off. We identify the point at which the camera cannot detect any further brightness and repeat this process for the selected camera sensors using their maximum parameters (**Table 2**). By measuring the LED’s total optical power, we can then calculate the theoretical minimal optical power that each camera was still able to detect in our setup. This calculation accounts for the respective sum of stacked optical densities,

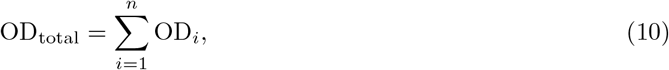

which yields the overall transmittance of the ND filters as,

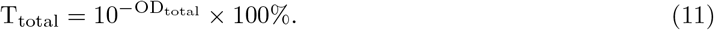

The theoretical minimal detectable optical power is obtained with P_*min*_ = T_total_ *×* P_LED_, assuming all other losses are negligible and summarized in (**Table 2**).

### Calculation of signal similarity with respect to the well size

For an interpretable quantification of the similarity of intensity distributions from distinct well sizes (image patches) we use the Jensen-Shannon divergence (JSD). From the signal values in a square region ℐ_*S*_, we construct a discrete probability distribution *P* = {*p*_*i*_*}* by computing a normalized histogram of pixel intensities over uniform bins (n = 15).

Given two such distributions *P* = {*p*_*i*_*}* and *Q* = {*q*_*i*_*}*, representing signal value distributions from two different image regions, the JSD is defined as

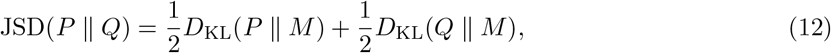

where,

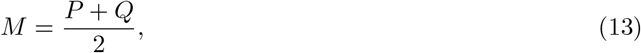

is the mixture distribution and *D*_KL_(· ∥ ·) is the Kullback-Leibler divergence:

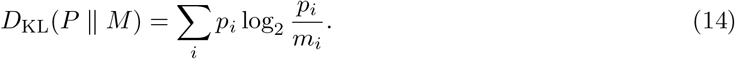

Using the base-2 logarithm ensures that the JSD is bounded between 0 and 1, where 0 indicates identical distributions and values closer to 1 indicate greater dissimilarity.

### Reagent preparation for chemiluminescent assays

Reagents **Table 5**, Materials **Table 6** and, Antibodies **Table 7** are listed in detail in the supporting information. Patient samples were acquired from Labcorp sample request SR2848 (see **Data Availability**).

The assay buffer was prepared as a 1x PBS solution containing 1% w/v BSA, 0.05% v/v Tween-20, 0.05% v/v ProClin300. After mixing, the solution was filtered through Sterile Filter Unit 0.22 µm and stored at 4 4 ◦C.

The mastermix diluent was made up containing 80% v/v HAMA Blocker, 10% v/v HRP-Stabilizer, 10% v/v Assay buffer, and stored at 4 ◦C.

To prepare the magnetic microparticle solution, 100 µL of Dynabeads M-280 Streptavidin (stock at 10 mg mL^−1^) was washed 3 times in assay buffer (3x 900 µL) by centrifuging the microparticles for 1 min at 12000 rpm removing the supernatant with a pipette and re-suspending in fresh assay buffer. Finally, the microparticles were diluted to a working concentration of 1 mg mL^−1^ in assay buffer.

### HRP- and Biotin-coupling of antibodies

For HRP-coupling, the conjugation of Anti-Renin Antibody 10850 with HRP was achieved using the EZ-Link™ Plus Activated Peroxidase Kit. The coupling is performed according to the manufacturer’s instructions. Antibodies were buffer exchanged using Vivaspin® 500 Centrifugal concentrator, Membrane 30000 MWCO PES (3x 500 µL PBS) and the resulting concentration determined using a NanoDrop Spectrophotometer 2000c prior to coupling and after HPLC-SEC purification.

Size exclusion chromatography (SEC) experiments were performed on an Agilent Bioinert 1260 HPLC system and an AdvanceBio SEC column (2.7 µm, 300 mm × 4.6 mm, 300 Å). The mobile phase was 150 mM sodium dihydrogen phosphate buffer, pH 7.0 (adjusted with sodium hydroxide). 50 µL samples and 5 µL molecular weight standards (Bio-Rad #1511901) were injected. The flow rate was set to 0.3 mL min^−1^ with a runtime of 25 min. Peaks were detected at 230, 260, and 280 nm. 10 runs were performed and the HRP coupled antibody fractions were combined.

For the Biotin-coupling, the conjugation of Anti-Aldosterone Antibody HM783 and Anti-Renin Antibody 10852 with biotin was achieved using EZ-Link NHS-PEG4-Biotin. Antibodies were buffer exchanged using Vivaspin® 500 Centrifugal concentrator, Membrane 30000 MWCO PES (3x 500 µL PBS), and the resulting concentration determined using a NanoDrop Spectrophotometer 2000c prior to coupling and after the coupling reaction. A 20-fold molar excess of biotin reagent (at 20 mM in DMSO) was added to the buffer-exchanged antibody, and the reaction was incubated for 2 h on ice. After the final buffer exchange, the coupled antibody was diluted to a concentration of approx. 1 mg mL^−1^ using 1x PBS. Modified antibodies were stored at 4 ◦C. We recommend for longer-term storage (> 1 week) to store affinity reagents directly in the mastermix diluent.

### Renin and Aldosterone Mastermix formulations

HRP-coupled anti-renin 10850 antibody was diluted to 0.98 µg mL^−1^ and the biotin-coupled anti-renin 10852 antibody to 0.89 µg mL^−1^ in mastermix diluent (RMM). The biotin-coupled anti-aldosterone antibody was stored separately from the HRP-adosterone to improve assay sensitivity. These two components of the mastermix were only mixed directly in the well after the sample was added. The two master mixes were: Aldosterone-3-CMO-HRP-Enzyme Conjugate at a 1:16,666 dilution in mastermix diluent (AMM1) or Biotin-Anti-Aldosterone Antibody HM783 at 0.88 µg mL^−1^ in mastermix diluent (AMM2).

### Bead Transfer Tool

To facilitate the manipulation of the magnetic microparticles used in the CLIA and to promote the use of readily available laboratory consumables, circumventing the cost of commercial alternatives, we developed a partially 3D-printed transfer tool (**Fig. 3b, Fig. Suppl. Fig. 7)**. The transfer tool consists of a plastic base with 2 mm holes spaced to match the pitch of the wells of a 96-well microplate **Fig. 3b**. In these holes 20 mm M2 bolts are screwed in. The steel bolts are then topped with 2 x 4 mm or 1 x 5 mm long, 2 mm diameter neodymium magnets. The holder with magnets attached is then placed into 8-well PCR strips, and the whole assembly (with the PCR strips pointing down so that their outside touches the liquid) is placed into the 96-well microplate at the desired position (see usage **Fig. Suppl. Fig. 7a-f**). The magnets attract the microparticles to the outer plastic walls, and the whole assembly can then be lifted out and placed in a new position on the 96-well microplate. To release the particles, the magnet holder and magnets are removed from the PCR strip, and the PCR tubes are agitated in the wells to disperse the particles (**Suppl. Fig. 7e-f**). CAD files are included in our data repository (see **Data Availability**).

### Assay Method

50 µL of sample was added to the well of a 96-well assay plate. Then 50 µL RMM (for renin) or 20 µL of AMM1 and 50 µL of AMM2 (for aldosterone) of the mastermixes were added to each sample. The reaction was incubated for 20 min at 500 rpm on an IKA MS3 digital shaker at r.t., protecting the plate from light exposure. After incubation 50 µL of the magnetic microparticle solution was added to separate wells. The microparticles were collected with the bead transfer tool and transferred to wells holding the sample and master mix. The reaction was then incubated for 10 min at 500 rpm at r.t., protecting the plate from light exposure. The microparticles were washed by moving them three times to three separate wells each containing 100 µL of assay buffer using the magnetic handling tool.

### Chemiluminescent Readout

The Luminata™ Crescendo imaging solution was prepared according to the manufacturer’s instructions. We added 100 µL of the imaging solution per well into white well C8 strips. Following the 3 washing steps the magnetic beads were transferred to the substrate using the magnetic handling tool. We measured luminescence using either our mobile reader (MR) with various image acquisition settings or a Bioteck Synergy H1 microplate reader (PR) with 1 s exposure and 60 s of shaking prior to reading and set the temperature at 28 ◦C.

### Standard curve samples

Renin standard curve samples: Recombinant human renin protein was diluted in renin/prorenin (REN/ PREN) double depleted human plasma. Resulting concentrations: **Fig. 5a** (10000, 1000, 125, 32.25, 7.8 pg mL^−1^), **Fig. 6a** (5000, 1000, 750, 500, 250, 125, 75, 50, 25, 10, 5 pg mL^−1^, and blank). Aldosterone standard curve samples: Aldosterone was diluted to 10 ng mL^−1^ in methanol. Dilutions were made from this using human serum as a diluent. Resulting concentrations **Fig. 5b**: (2000, 500, 125, 32.25, 7.8 pg mL^−1^), **Fig. Suppl. Fig. 13:** (2000, 1000, 800, 600, 400, 200, 150, 125, 100, 50, 25 pg mL^−1^, and blank).

### Measurement of human plasma-EDTA and serum samples

Samples were obtained from Labcorp. The biospecimen type was matched Serum and EDTA Plasma from whole blood. The disease status of patients is unknown, but the patients were referred for a Labcorp Aldosterone:Renin Ratio test (test number: 004354). The clinical characteristics are unknown. Only age, gender, collection date, Renin Activity, Plasma Aldosterone Concentration and Aldosterone:Renin Ratio were provided. Samples were shipped at -78.5 ◦C on Dry Ice and stored at -80 ◦C except during analysis. The storage duration was less than 6 months. We analyzed 50 paired plasma-EDTA and serum samples from Labcorp encompassing a range of renin and aldosterone concentrations (**Table 3**). We measured aldosterone and renin concentrations using our developed CLIAs, measuring all samples with the SOTA PR and our MR. Images were taken in a series of six images with a 5s exposure and at ISO 1600. All samples were assayed undiluted and in technical triplicate. Aldosterone concentrations were measured in serum samples, and renin concentrations were measured in the paired plasma-EDTA samples, as done by the SOTA measurement by Labcorp. Labcorp values were converted from renin plasma activity (PRA; ng mL^−1^ h^−1^) to direct renin concentration (DRC; pg mL^−1^) using a conversion factor of 7.6 found in literature^47^. Labcorp aldosterone concentrations (ng dL^−1^) were converted with a factor of 10 to pg mL^−1^.

### Determination of assay signal saturation time

To identify the onset of signal saturation over time, we analyzed the smoothed signal trajectory 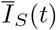 for each experimental condition (i.e., assay and concentration, *Sig*.). The signal was first smoothed using a moving average with a window of *w* time points. The first derivative *D*(*t*) of the smoothed signal was computed to evaluate changes in signal values (*d(Sig.)*). A dynamic saturation threshold *τ* was defined as a fixed fraction *f* of the maximum absolute derivative within the time series:

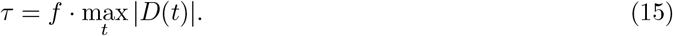

The signal was considered saturated when the absolute value of the derivative remained below *τ* for at least *d* consecutive time points. The saturation onset time *t*_*s*_ was defined as the first time point where this criterion was met:

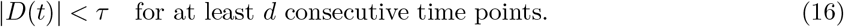

Parameter values used were *w* = 5 (smoothing window size), *f* = 0.5 (threshold factor), and *d* = 3 (minimum saturation duration time points).

### Modeling and statistical analysis

To model the relationship between signal and concentration, we fitted three candidate models: a linear model (LM), a four-parameter logistic model (4PL), and a five-parameter logistic model (5PL). LMs were fitted to the data in the form of

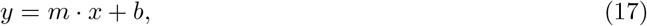

where, *y* is the predicted response, *m* the slope of the line and *b* the y-axis intercept and 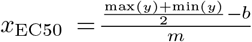. For the 4PL model

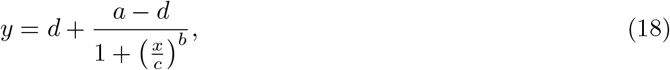

where *y* is the predicted response, *a* the minimum (maximum for decreasing curve respectively) asymptote, *d* maximum (minimum for decreasing curve respectively) asymptote, *c* the EC50 and *b* the Hill slope and *x*_EC50_ = *c*. To determine the concentrations (*x*) for a quantified signal (*y*) we use 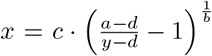. For the 5PL model taking an asymmetry into account

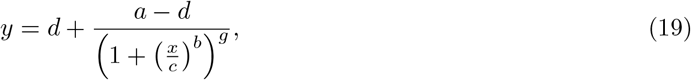

where, *g* is the asymmetry factor. To determine the concentration (*x*) value, we solve the 5PL model numerically as

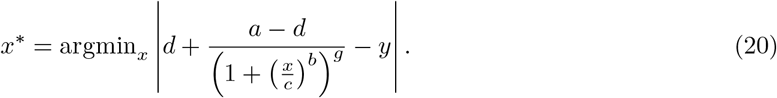

Model selection was based on two criteria: (i) the lowest corrected Akaike Information Criterion (AICc), which adjusts Akaike’s Information Criterion (AIC) for finite sample sizes, and (ii) a high coefficient of determination (*R*^2^). For each model, AICc was computed as:

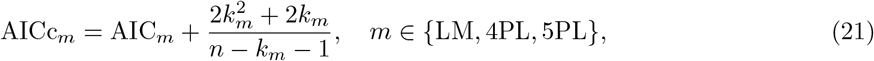

where, (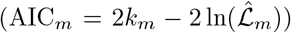, (*k*_*m*_) is the number of parameters in model (*m*), (*n*) is the number of observations, and 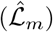 is the maximum likelihood of the model. For each model, the relative quality was assessed using ΔAICc, defined as:

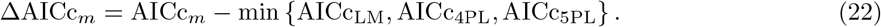

The model with (ΔAICc_*m*_ = 0) was considered the simpler model. We use ΔAICc_*m*_ = 0 as the primary model selection criterion. Models with (ΔAICc_*m*_ *<* 4) are considered to have comparable statistical support. The final selected model was the one with the highest (*R*^2^). For a few cases, the 4PL model showed the lowest AICc, with the 5PL ΔAICc ranging from 2.39 to 8.21 (**Table 12**). To ensure consistent interpretation and account for potential response asymmetry, the 5PL was used across all datasets.

### Performance study sample size considerations

In a pilot study (*n* = 12; six patient samples measured in duplicate), our renin assay demonstrated a strong correlation with the independent Labcorp reference method (*R*^2^ = 0.869; Pearson *r* = 0.93, 95% CI: 0.77–0.98; *p* = 9.98 *×* 10^−6^). Although encouraging, the confidence interval was wide, reflecting the inherent imprecision of small pilot studies. To plan the subsequent validation study, we calculated the required sample size using the Fisher *z*-transformation of the correlation coefficient:

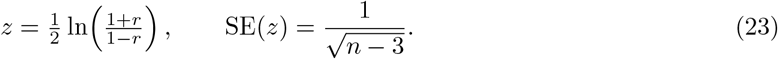

The 95% confidence interval (CI) for *z* is defined as

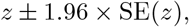

which is then back-transformed to the correlation scale. Assuming a true correlation of *r* = 0.90, a sample of approximately *n* = 50 observations yields a 95% CI width of about 0.11, providing sufficient precision to confirm the strength of association observed in the pilot. Based on this calculation, we designed the performance study with 50 independent human samples (each measured in triplicate).

### Mixed effects modeling

To quantify systematic bias relative to an independent LC/MS reference (Labcorp) and to assess device interchangeability between the mobile reader (MR) and the plate reader (PR), we employed linear mixed-effects models accounting for repeated technical measurements per sample.

Bias was defined as the difference between the assay-derived concentration and the LC/MS reference (both in pg/mL):

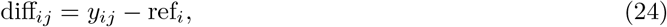

where *y*_*ij*_ denotes the concentration measured by device *j* (MR or PR) for sample *i*, and ref_*i*_ denotes the corresponding LC/MS reference concentration. Assay readouts were converted to concentrations using assay-specific standard curves. For renin PRA LC/MS values, a published conversion factor of 7.6 was first applied (**Table 4**).

We modeled bias as:

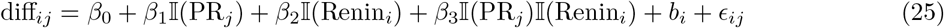

where *β*_0_ represents the mean bias for MR aldosterone measurements, *β*_1_ the device effect (PR vs MR) for aldosterone, *β*_2_ the assay effect (renin vs aldosterone) for MR, and *β*_3_ the interaction between device and assay. The random intercept 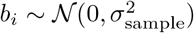) captures sample-level variability, and *ϵ*_*ij*_ ∼ 𝒩 (0, *σ*^2^) denotes residual error.

Planned comparisons (a priori contrasts) were used to test absolute bias relative to zero for each device–assay combination, and device differences within each assay.

To assess proportional agreement with the LC/MS reference (Labcorp), concentrations were log-transformed and modeled as:

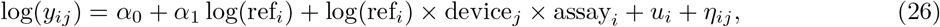

where *α*_1_ represents the calibration slope for MR aldosterone measurements, interaction terms allow slopes to vary by device and assay, and *u*_*i*_ ∼ 𝒩 (0, *τ*^2^) denotes a sample-level random intercept. Slopes were interpreted as calibration factors, with a slope of 1 indicating proportional agreement.

The relative contribution of sample-level variability was quantified using the intraclass correlation coefficient (ICC), defined as:

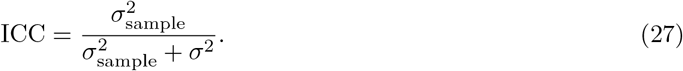

### 4.1 AI Assistance Disclosure

Generative AI tools integrated into Overleaf and Perplexity were used for language refinement and for drafting parts of the analysis and visualization code. All outputs were critically reviewed and validated by the authors.

## Data Availability

All data produced in the present study are available upon reasonable request to the authors.

## 5 Data and code availability

Data and analysis scripts underlying this study will be provided to the reviewers and deposited in a publicly accessible data repository upon publication. Additional data related to this paper may be requested from the authors upon reasonable request.

## 6 Acknowledgments

This work was supported by Innosuisse (57350.1 IP-LS). We would like to thank Christoph Schumacher, Teresa Gerlock, Ronald E. Steele, Damian Pharma AG, Therina Du Toit (Universitätsklinik für Nephrologie und Hypertonie Inselspital, Universitätsspital Bern), Daniela Tobler (Institut für Pharmatechnologie und Biotechnologie, School of Life Sciences, University of Applied Sciences and Arts Northwestern Switzerland) and Daniel Gygax for helpful discussions. We thank Jonathan Schmidli and Markus Klement (CRFB and mechanical workshop, Department of Biosystems Science and Engineering, ETH Zurich) for their help with microfabrication and high-resolution 3D printing. We thank Hans Michael Kaltenbach and Joerg Stelling for helpful feedback on the manuscript.

## 7 Author contributions

C.S.W. and A.P.C. conceived the study, generated the hypotheses, and designed the experiments with the help of S.A., J.A., A.G., and D.M.M,. C.S.W., S.A., J.A., A.G. and A.P.C. performed all the experiments and analyzed the data. A.P.C wrote the data analysis scripts. C.S.W., S.A., J.A., D.M.M and A.P.C. analyzed and interpreted data. J.A. and A.P.C. conceived and designed the smartphone app. A.P.C. conceived and designed the MCCS and MR. C.S.W., S.A. conceived and designed the CLIA assays and transfer tool. D.M.M and A.P.C. jointly supervised the project. C.S.W., S.A., J.A. and A.P.C. wrote the manuscript. All authors contributed to manuscript review, revision, and finalization.

## 8 Competing interests

A.P.C. is an inventor on a provisional patent application related to this work filed by ETH Zurich (EP25197418). The authors declare that they have no other competing interests.

## Notes

### Author Declarations

Ethikkommision Nordwest- und Zentralschweiz (EKNZ) waived ethical approval for this work.

