## Supporting Information for "_CLIA_MDK: A Modular Smartphone Platform Matching Plate Reader Performance for Chemiluminescent Immunoassay Development"

### Supplementary Information

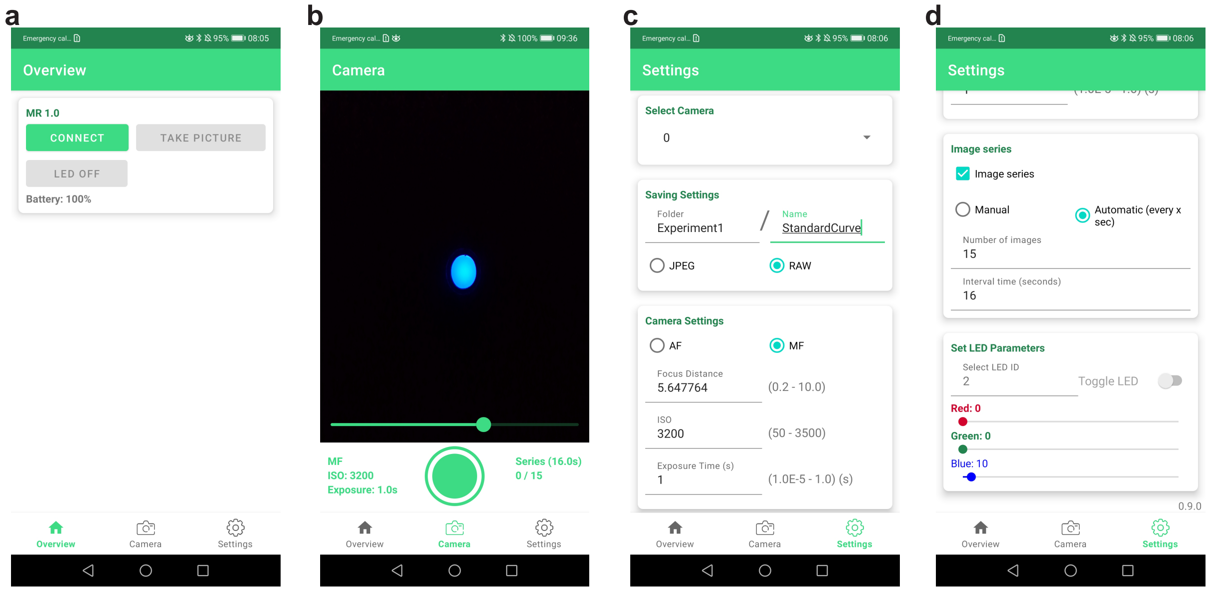


Suppl. Fig. 1: App functionalities. **a** Overview view; allows a connection via Bluetooth with the MCCS, and when connected, to capture an image or switch the LEDs on and off. **b** Camera view; Allows to set a focus and start the image acquisition manually. **c** Settings view. The 'Select Camera' panel allows to choose among available camera sensors. The 'Saving Settings' allows to specify a folder and an image name as well as to select the image format between compressed (JPG) and uncompressed (RAW). Within 'Camera Settings', the focus mode, auto focus (AF) or manual focus (MF) is set. For reproducibility, the focus motor position can be saved and set using actual numbers between experiment series. Further, the ISO and Exposure time in seconds are set. Some camera models allow the use of values outside the automatically detected range. **d** The 'Image series' can be used in manual or automatic mode to take a series of subsequent images manually or automatically, respectively for a defined number of images with a constant interval in seconds. The 'Set LED Parameters' allows control of the MCCS LEDs by ID and red, green, and blue brightness within an 8-bit range.

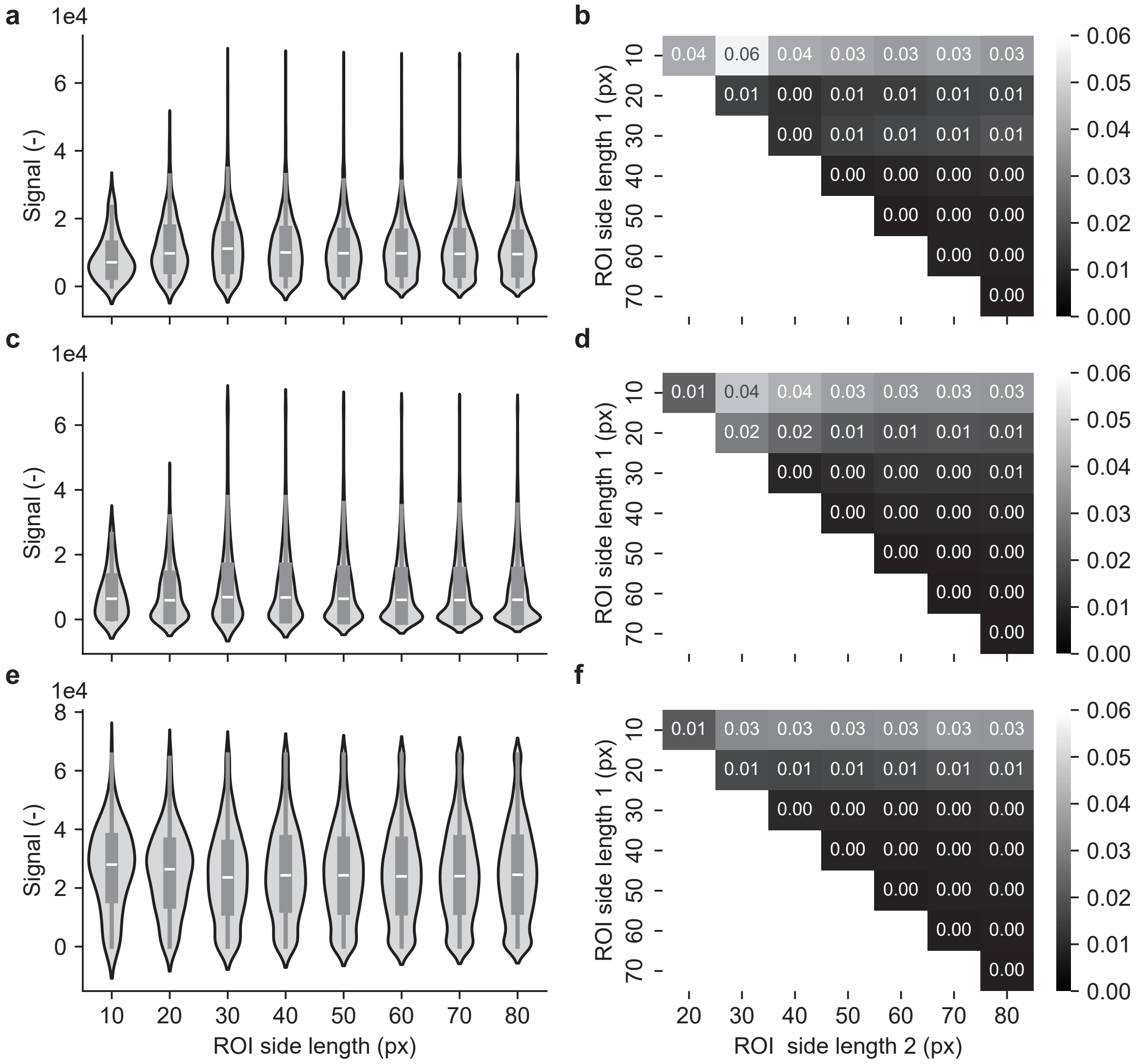


Suppl. Fig. 2: Effect of the square region of interest (ROI) side length on signal mean. **a** Violin plots of the raw pixel value distribution plotted against square ROI side length in pixels, resulting in $n = 121, 441, 961, 1681, 2601, 3721, 5041, 6561px$ considered on an image taken with the MCCS and X13P with $8 \cdot 0.9$ ND filters and the LED's blue channel set to level 10 ($8.9 \pm 0.03 \times 10^{-15}$ W). **b** Similarity in the probability distribution quantified by the Jensen–Shannon divergence (JSD) with bin size = 15 of the pairwise comparison between the square ROI side lengths. JSD ranges between 0 (identical) and 1 (maximally different); values $< 0.02$ indicate negligible distribution differences, and any square ROI side length $> 10$ pixels can be used. **c, d** same as **a,b** but for a CLIA with a relatively low signal for 125 pg mL$^{-1}$ renin. **e, f** same as **a,b** but for a CLIA with a relatively medium signal for 500 pg mL$^{-1}$ renin.

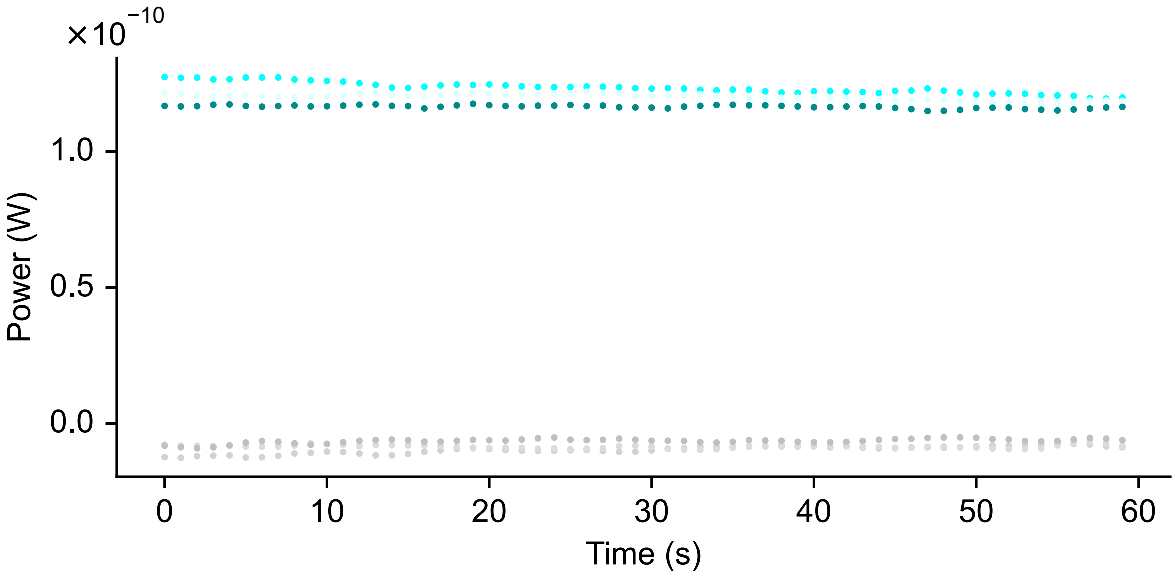


Suppl. Fig. 3: Power measurements of an aldosterone sample (100 $\text{pg}\,\text{mL}^{-1}$) in a white well-strip well and measurement of an empty well over time ($n = 3$). On average, the power measured for aldosterone is 1.20e-10 ± 3.18e-12 W (mean ± SD) and for the empty well, -8.19e-12 ± 1.68e-12 W (mean ± SD).

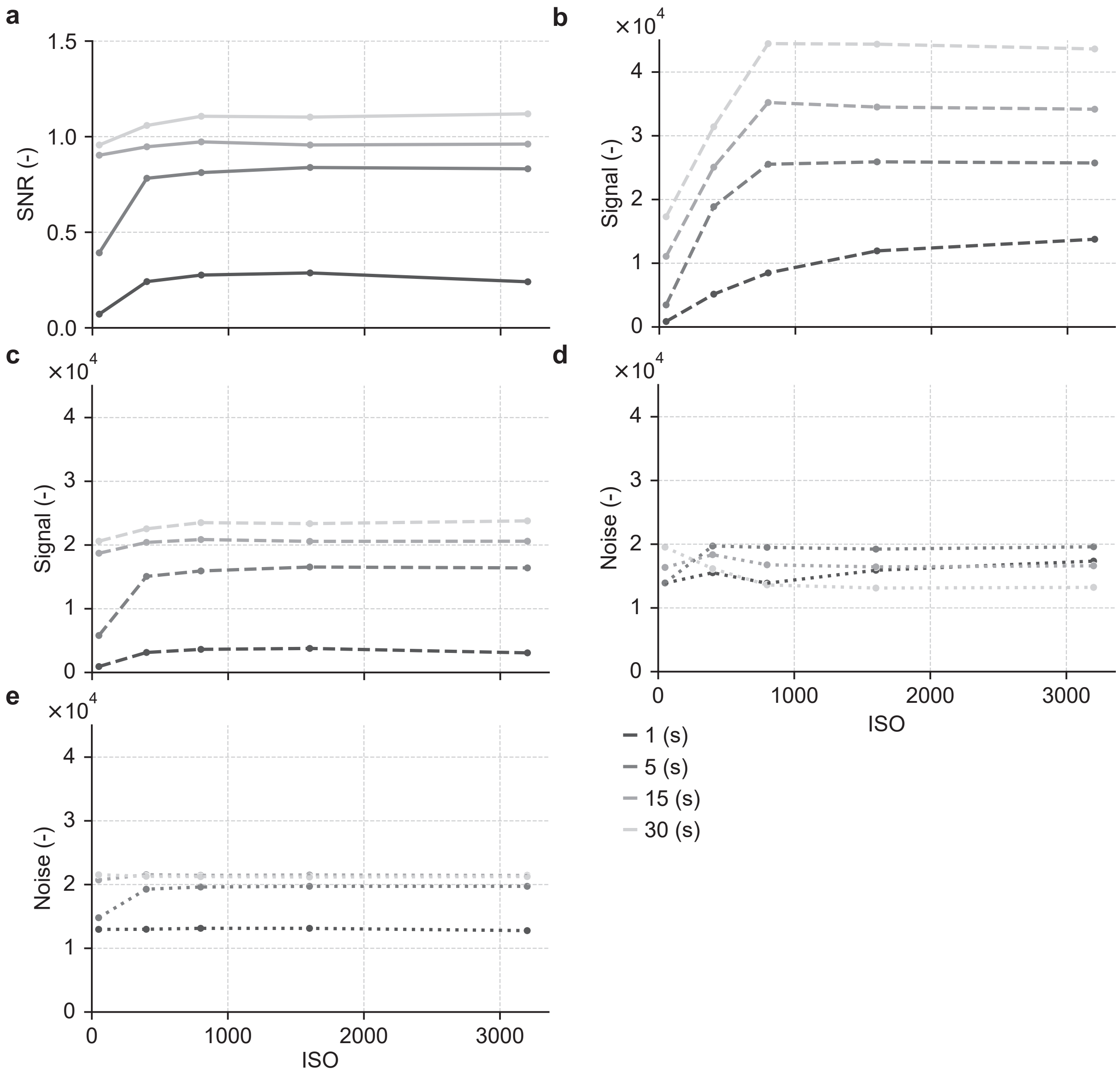


Suppl. Fig. 4: Signal, noise and signal-to-noise ratio (SNR) as a function of ISO and exposure time. **a** SNR for the HP20 camera sensor (grey dots) for different exposure times (1, 5, 15, and 30 s) and ISO (50, 400, 800, 1600, 3200) with visual guides indicating the trend (grey lines). **b,c** Signal response (grey dots), same conditions, with visual guides (grey dashed lines) measured with the X13P and HP20, respectively. **d, e** Noise (grey dots), same conditions, with visual guides (grey dotted lines) measured with the X13P and HP20, respectively.

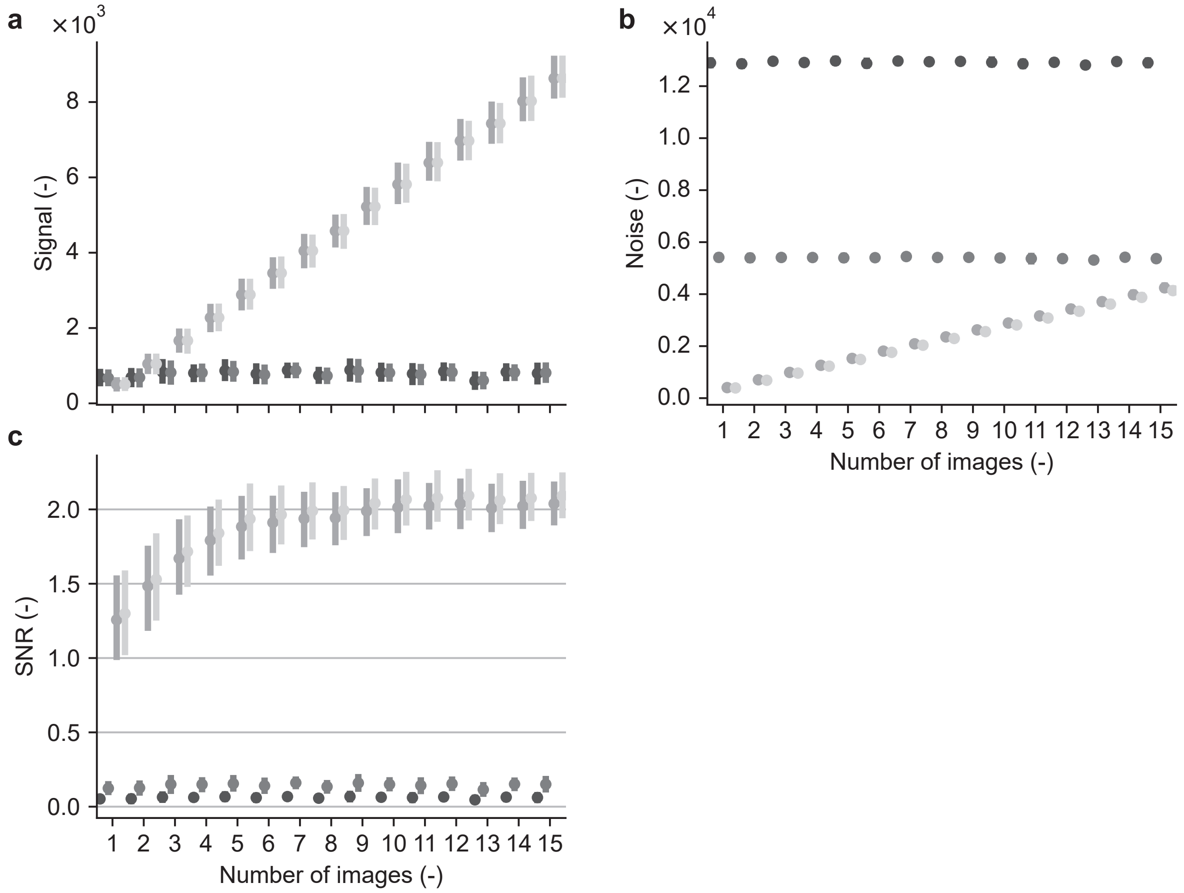


Suppl. Fig. 5: Signal, noise, and signal-to-noise ratio (SNR) as a function of the number of sequential images taken with X13P and signal processing. **a** Signal for different signal processing for RAW (dark grey) control for no-processing, ROF (medium dark grey), NREA (medium light grey) and $\mathrm{NREA}_{\mathrm{ROF}}$ (light grey). **b** same as in **a** for Noise. **b** same as in **a** for SNR

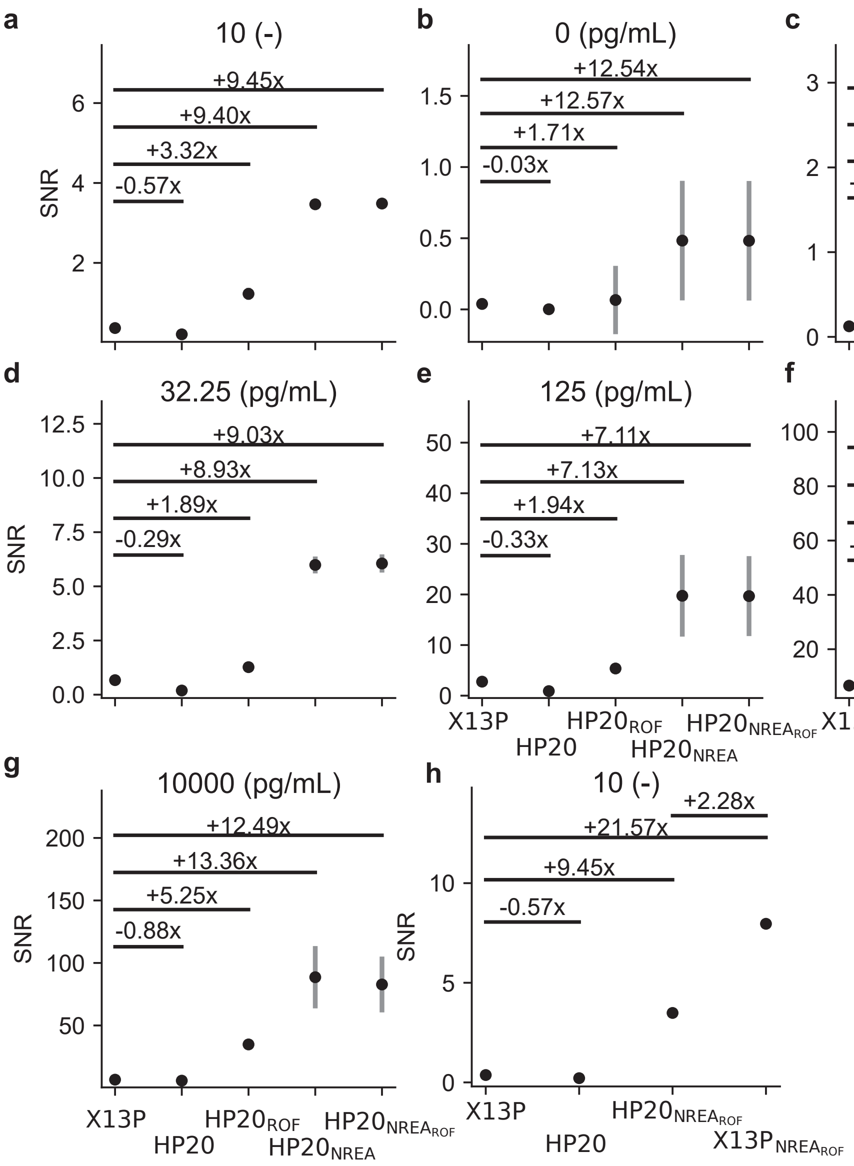


Suppl. Fig. 6: Signal-to-noise ratio (SNR) as a function of a recent smartphone camera sensor (X13P) and an older one (HP20, release 2018), and signal enhancement. **a** SNR for an *in silico* signal taken with the MCCS and the blue LED with intensity set to 10, ISO 3200, and 30 s. Data (black dots, vertical lines, mean $\pm$ SD) with fold change (x) compared to the X13P raw signal for the raw signal (HP20), the ROF denoised signal ($HP20_{ROF}$, 0.9), the NREA enhanced raw signal ($\mathrm{HP20_{NREA}}$), and the NREA enhanced ROF denoised signal ($\mathrm{HP20_{NREA_{ROF}}}$), each considering two images in series and two technical replicates. **b-g** Same as in **a** but for a renin standard curve using different concentrations $(0, 7.8, 32.25, 125, 10^3, 10^4$ $\mathrm{pg\,mL^{-1}}$ and ISO 3200, and 15 s. **h** SNR of RAW and $\mathrm{NREA_{ROF}}$ processed signals using the X13P and HP20. The algorithm improved the SNR of the HP20 9.45 folder relative to X13P (raw), demonstrating that older sensors can achieve comparable performance. However, modern sensors still exhibit higher photon sensitivity, as evidenced by the 2.28-fold increase in X13P relative to HP20 when the algorithm was applied to both.

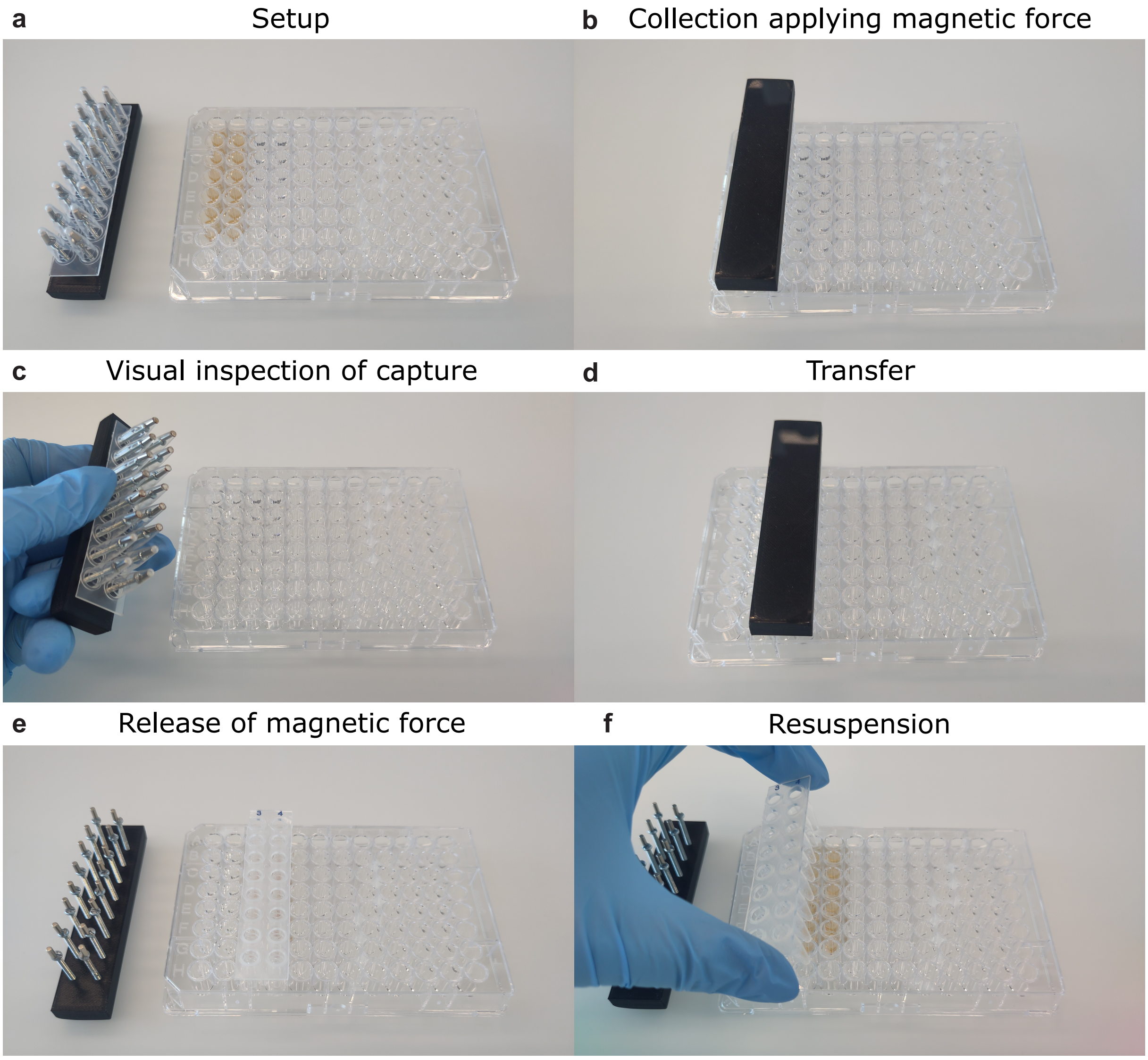


Suppl. Fig. 7: Magnetic microparticle transfer tool and its application. **a** In the initial setup, magnetic microparticles are in solution in a 96-well standard microplate. The magnetic holder is assembled and covered with PCR strips or plates cut to size to serve as tip combs. **b** Collection of magnetic microparticles by inserting the magnetic transfer tool into the magnetic microparticle solution. **c** After a few seconds of magnetic microparticle collection, the transfer tool is removed from the solution and visually inspected to confirm complete capture of microparticles. **d** Transfer to next solution (e.g. wash buffer or chemiluminescence substrate). **e** Release of magnetic force to release the magnetic particle from the tip comb. **f** Resuspension of particles in solution by gently moving the tip comb from side to side. Finally, the tip comb is removed from the solution. The size of the magnetic transfer tool can be scaled from 1-12 columns if desired. For CAD files and assembly instructions, see **Data Availability** section.

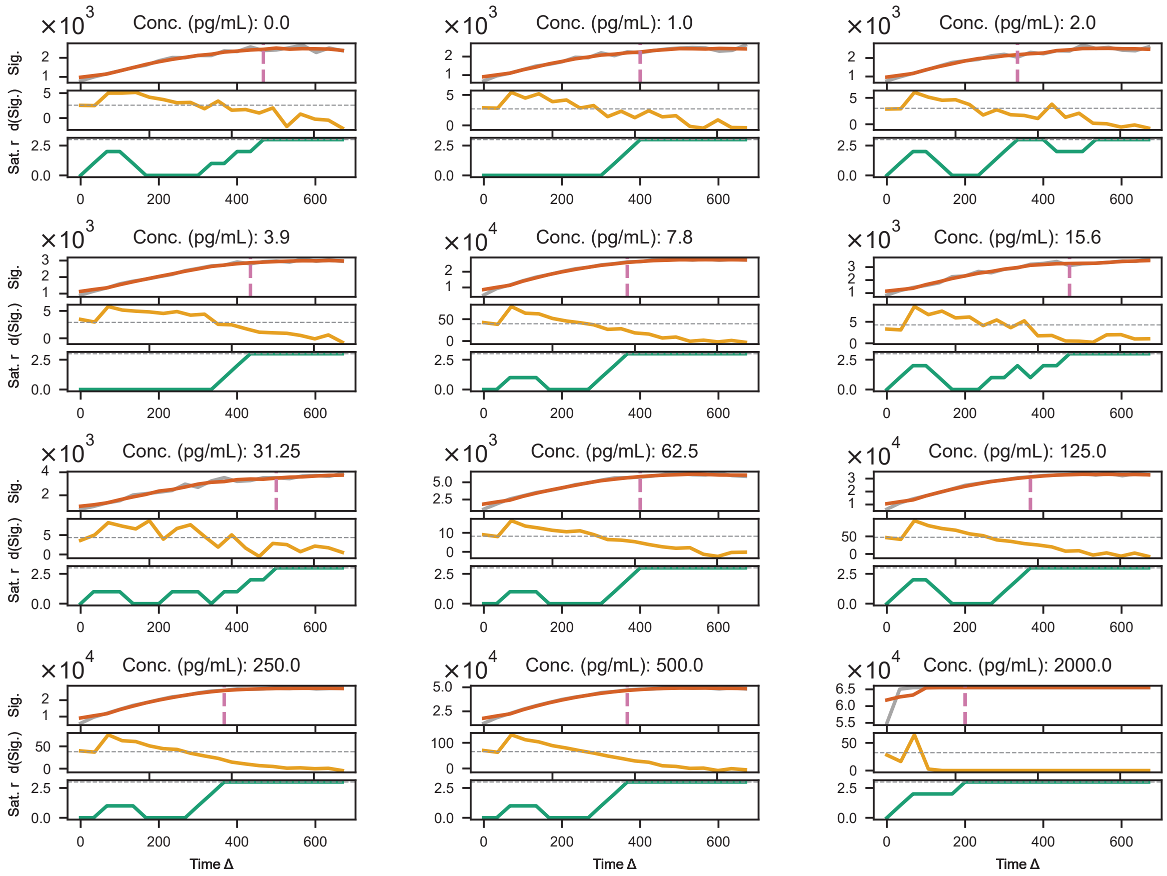


Suppl. Fig. 8: Quantification of assay signal stability for different assay concentrations as a function of time (in seconds, Δ Time). Renin assay signal (Sig.) (grey line) with smoothed signal (red line); vertical dashed line (pink) shows the estimated onset of signal saturation (top panel). First derivative of the smoothed signal (d(Sig.), orange), with horizontal dashed line (gray) indicating the saturation detection threshold (middle panel). Cumulative number of consecutive time steps meeting the saturation criterion (Sat. r. green); the horizontal dashed line (gray) indicates the minimum required streak for saturation assignment. The average time is $400s$.

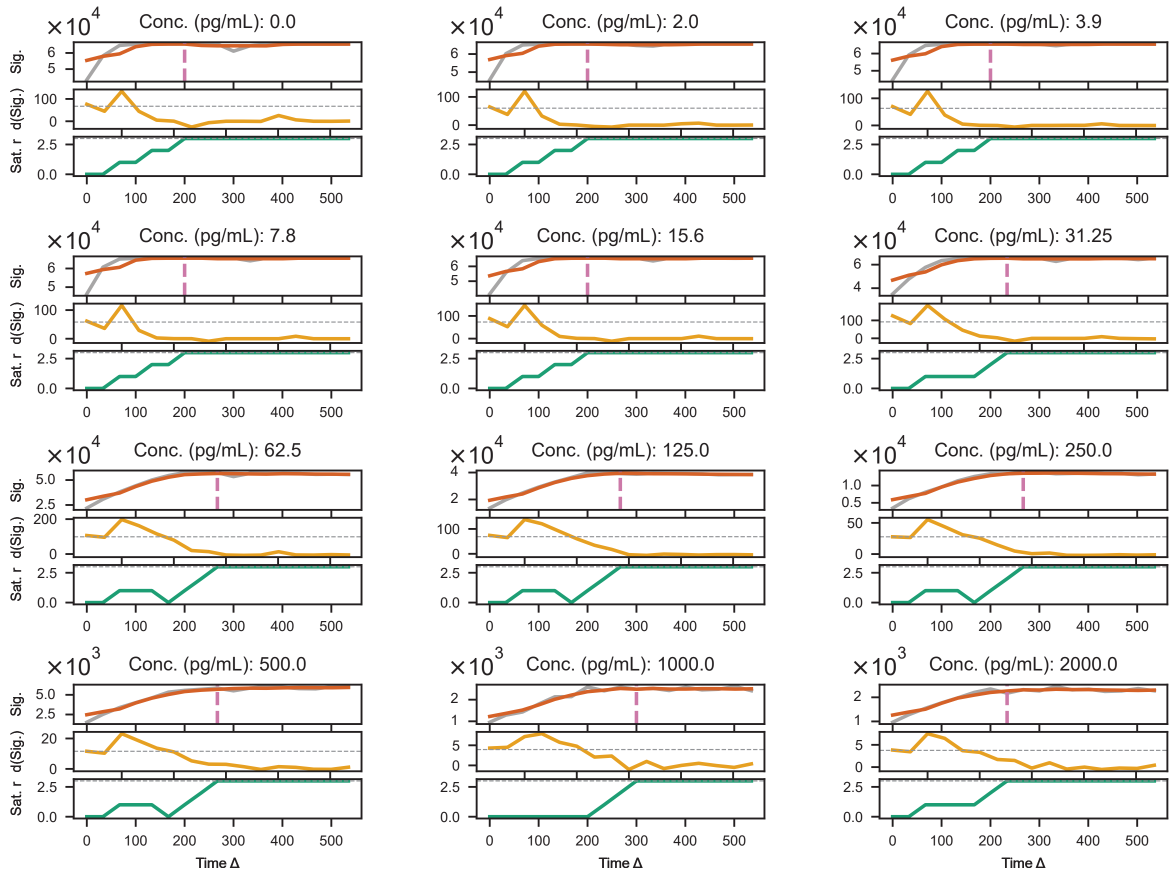


Suppl. Fig. 9: Quantification of assay signal stability for different assay concentrations as a function of time (in seconds, Δ Time). Aldosterone assay signal (Sig.) (grey line) with smoothed signal (red line); vertical dashed line (pink) shows the estimated onset of signal saturation (top panel). First derivative of the smoothed signal (d(Sig.), orange), with horizontal dashed line (gray) indicating the saturation detection threshold (middle panel). Cumulative number of consecutive time steps meeting the saturation criterion (Sat. r. green); the horizontal dashed line (gray) indicates the minimum required streak for saturation assignment. The average time is 200$s$.

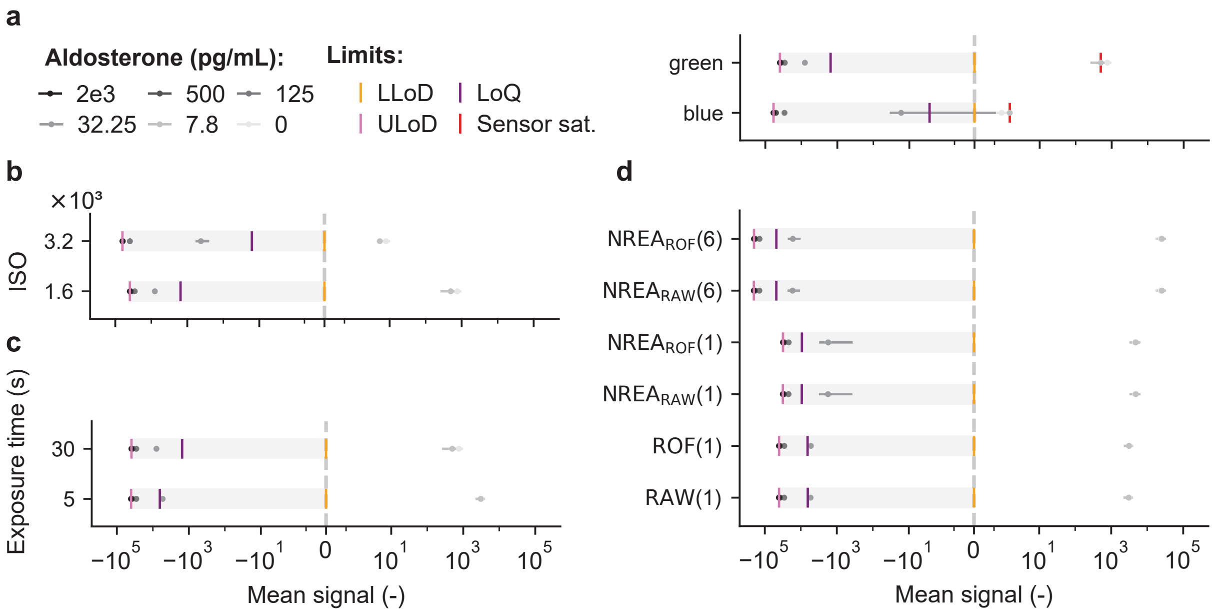


Suppl. Fig. 10: **a** Channel-based dynamic range for an aldosterone assay at different concentrations with the average signal (black dot, vertical line (mean ± SD), $n = 2$) normalized to the LLoD (vertical orange line) with limit of blank (vertical cyan line), limit of detection blank (vertical blue line), LoQ (vertical violet line), ULoD (vertical pink line) and sensor saturation (vertical dashed red line, at 16-bit). The grey box indicates the range between LLoD and ULoD. The further apart the data and limits, the larger the dynamic range. **b** Same as in **a** with the green ch and different ISO values (1600, 3200, 12800). **c** Same as in **a** with the green ch, ISO 1600 for different exposure times ($t_{exp}$ = 5, 15, 30 s). **d** Same as in **a** with the green ch, ISO=1600, $t_{exp}$ = 5 s for unprocessed (RAW), ROF denoising (ROF), NREA on RAW or ROF denoised images ($NREA_{RAW}$, $NREA_{ROF}$) for one and a series of six images.

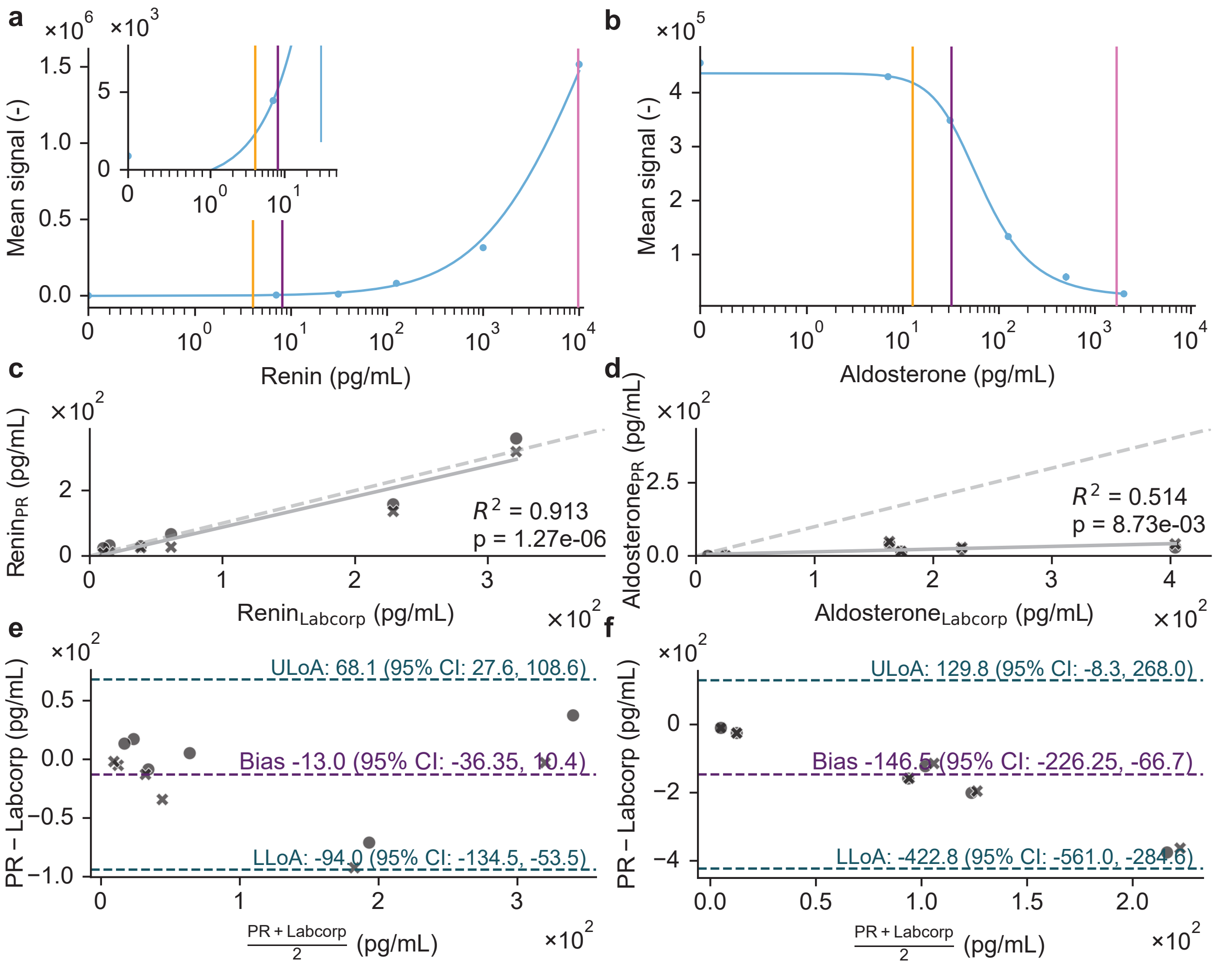


Suppl. Fig. 11: Our CLIA assay measured with SOTA PR in comparison with LC/MS SOTA Labcorp using patient samples. **a** Renin standard curve (blue dots and vertical lines, mean ± SD) with 5-parameter logistic regression (5PL) fit (blue line) recorded with PR ($n = 6$). The standard curve was measured using spiked renin/prorenin double-depleted plasma. **b** Aldosterone standard curve (blue dots and vertical lines, mean ± SD) with 5-parameter logistic regression (5PL) fit (blue line) recorded with MR with proposed signal processing ($n = 6$). The standard curve was measured in spiked human serum. **c, d** PR correlation with SOTA (Labcorp) measurements for renin and aldosterone, respectively ($n = 6$). **e, f** Bland-Altman plot comparing the PR results with Labcorp results for renin and aldosterone, respectively ($n = 6$).

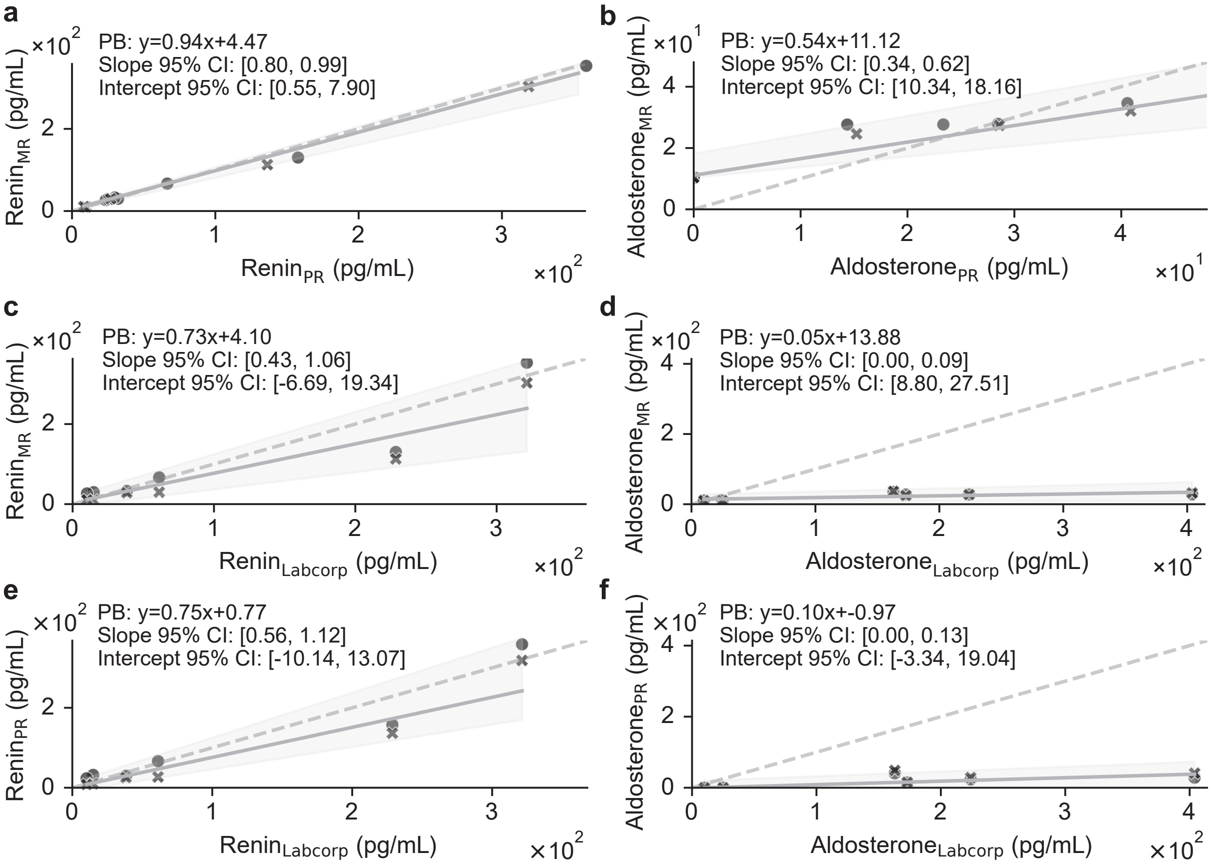


Suppl. Fig. 12: Method comparison with Passing–Bablok regression between MR and PR. **a** Patient samples ($n = 6$) for renin measured with MR and PR, Passing-Bablok regression (grey line), 95% CI (grey region), and identity line (grey dashed line). **b** same as in **a** for aldosterone. **c** same as in **a** but MR compared to SOTA (Labcorp). Labcorp values were converted from renin plasma activity (PRA; $\mathrm{ng\,mL^{-1}\,h^{-1}}$) to direct renin concentration (DRC; $\mathrm{pg\,mL^{-1}}$) using the conversion factor 7.6. **d** Same as in **c** for aldosterone; no conversion factor applied. **e,f** same as in **c,d** signals quantified with PR for renin and aldosterone, respectively.

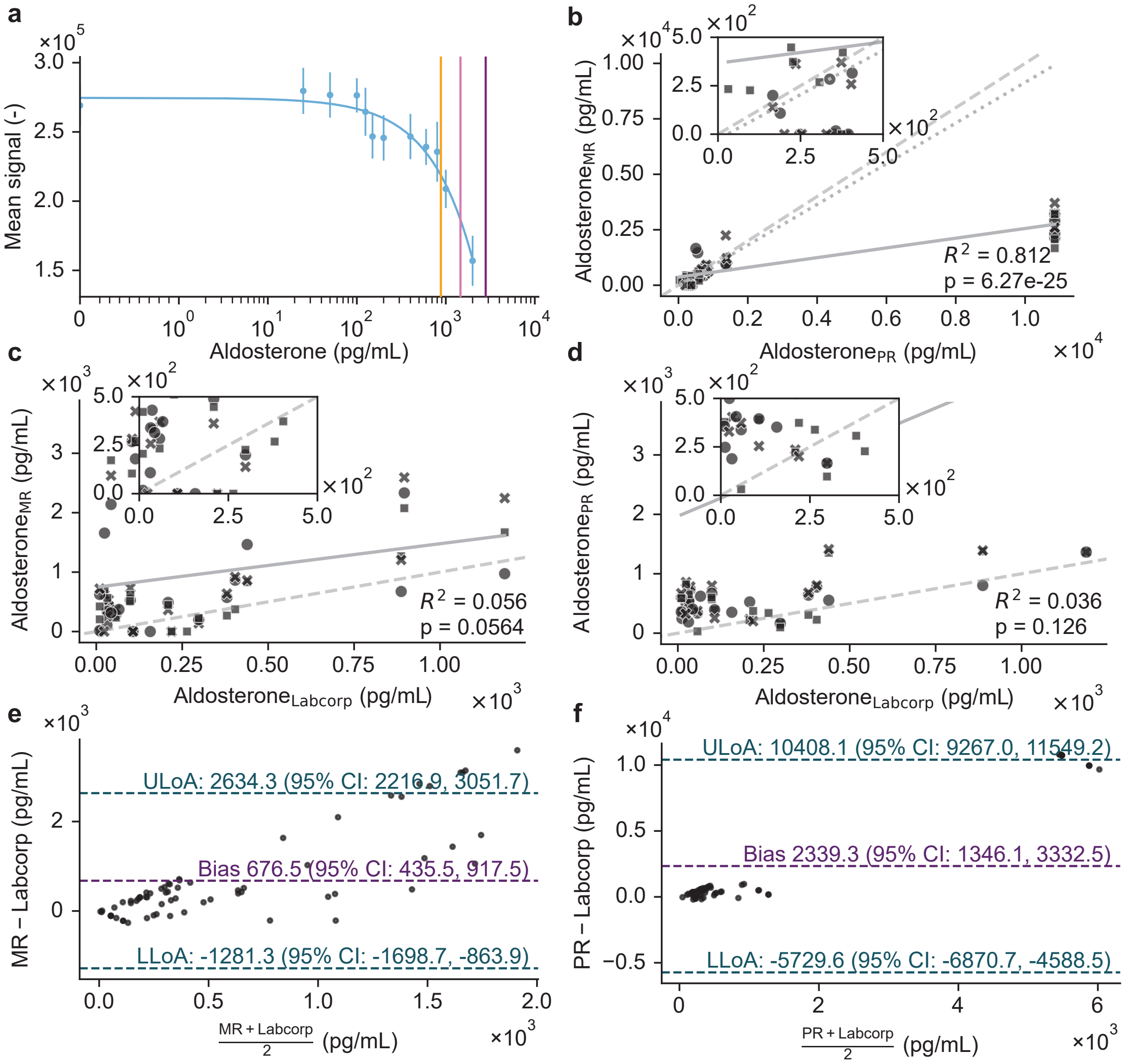


Suppl. Fig. 13: Performance study comparing MR with SOTA using patient plasma samples on the aldosterone assay. **a** Aldosterone standard curve with 5-parameter logistic regression (5PL) fit (blue line) and data (mean ± SD) recorded with MR with proposed signal processing ($n = 6$). The standard curve was measured using spiked serum. Vertical lines show LoD (yellow), LoQ (purple), ULoD (pink). **b** Pearson correlation (grey line) of MR to PR measurements of patient plasma samples ($n = 50$). **c** Pearson correlation (grey line) of MR aldosterone quantification with SOTA (Labcorp) measurements for plasma samples (n=50). Labcorp values were converted from renin plasma activity (PRA; $\mathrm{ng\,mL^{-1}\,h^{-1}}$) to direct renin concentration (DRC; $\mathrm{pg\,mL^{-1}}$) using the conversion factor 7.6. **d** Pearson correlation (grey line) of PR aldosterone quantification with Labcorp measurements ($n = 50$). **e, f** Bland-Altman plot comparing the results with Labcorp for MR and PR quantification, respectively ($n = 50$).

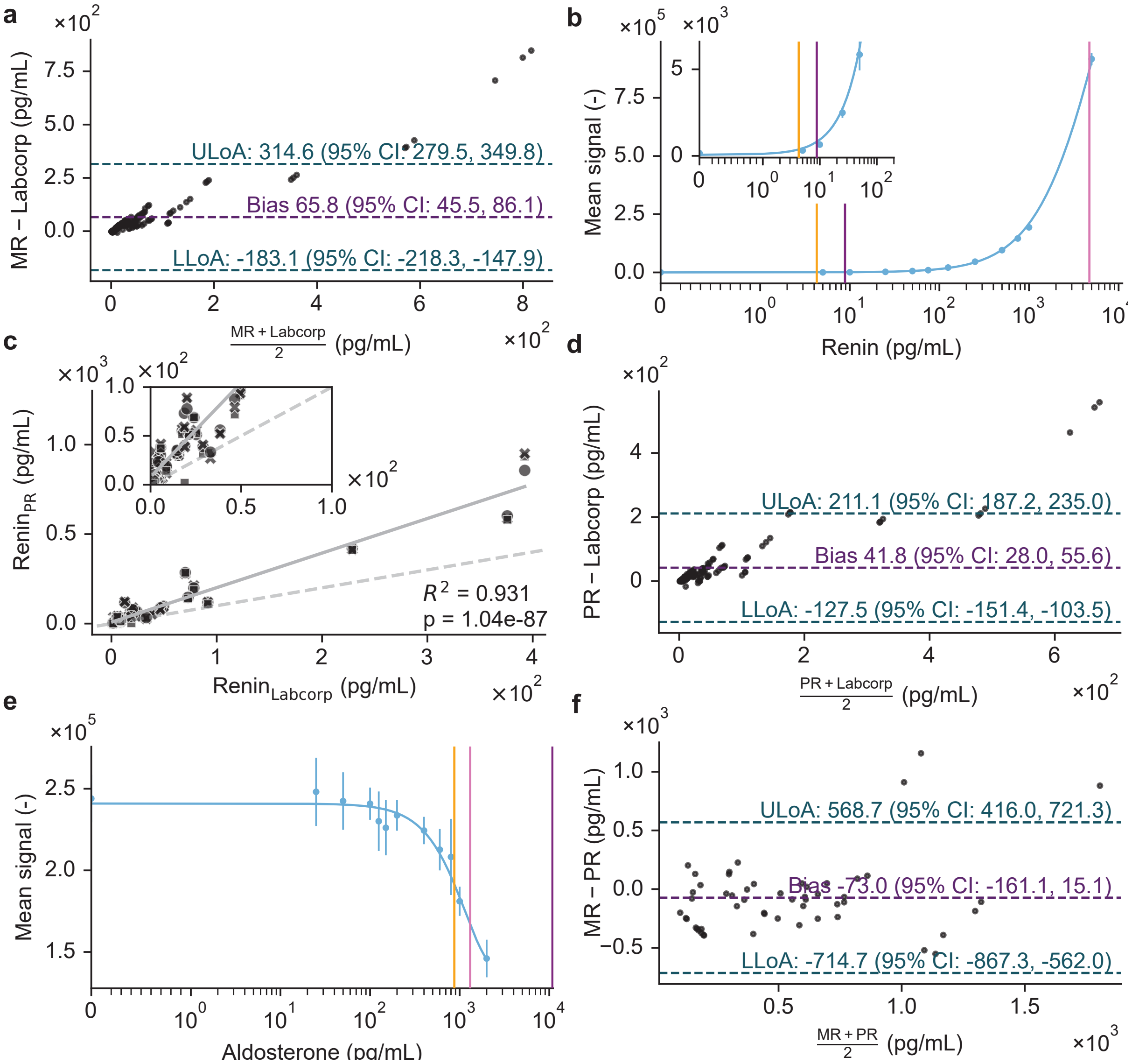


Suppl. Fig. 14: PR standard curves and comparison to MR. **a** Bland-Altman plot comparing the MR results with Labcorp ($n = 50$). **b** Renin standard curve with 5-parameter logistic regression (5PL) fit (blue line) and data (mean ± SD) recorded with PR ($n = 6$). The standard curve was measured using spiked rein/prorenin double-depleted plasma. Vertical lines show LoD (yellow), LoQ (purple), ULoD (pink). **c** Correlation of PR renin quantification with an independent method (LC/MS, Labcorp) for plasma samples ($n = 50$). **d** Bland-Altman plot comparing the PR results with Labcorp ($n = 50$). **e** same as in **b** for an aldosterone standard curve in spiked human serum. **f** Bland-Altman plot comparing the MR results with PR quantification, respectively ($n = 50$).

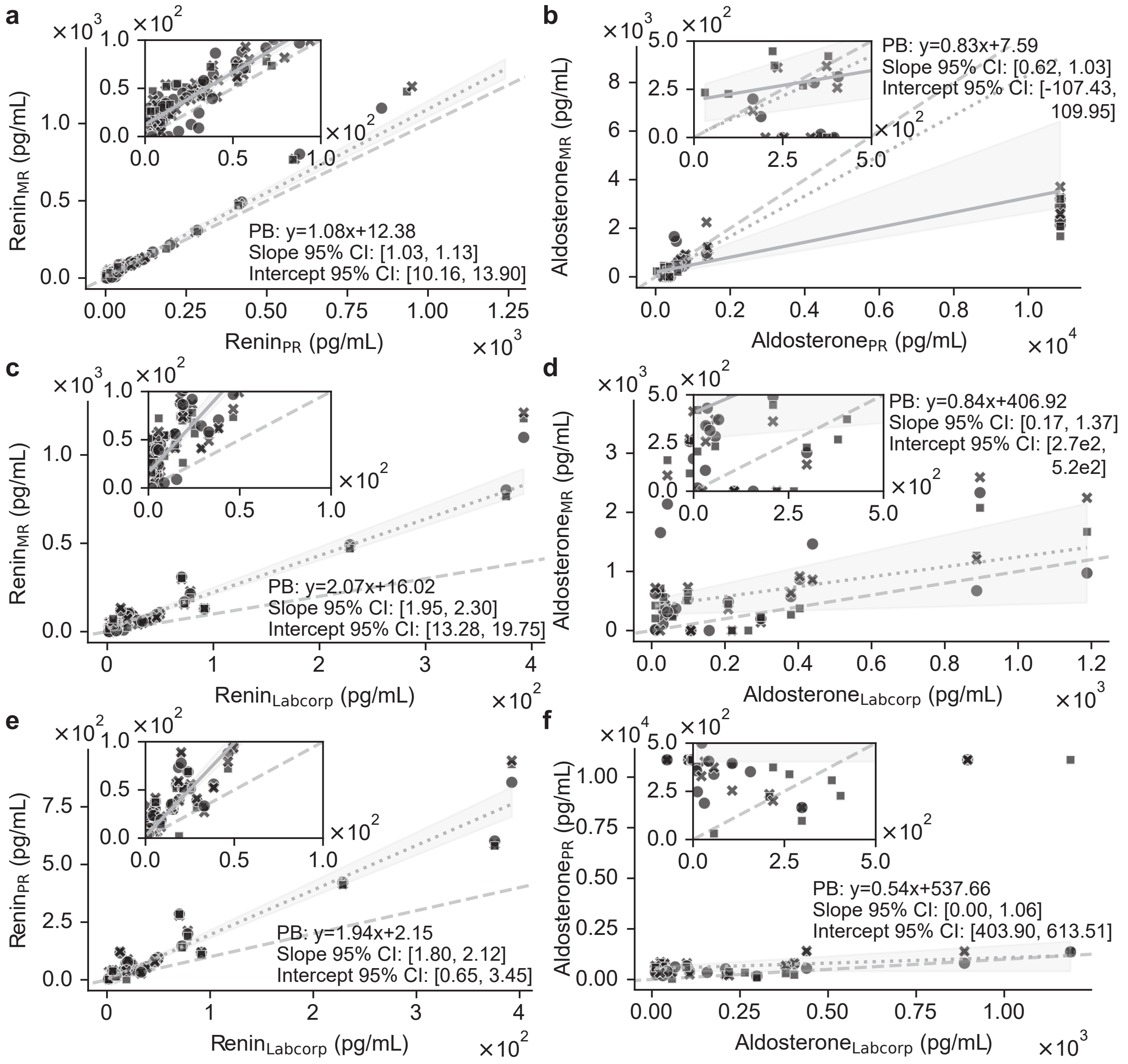


Suppl. Fig. 15: Method comparison with Passing–Bablok regression between MR and PR for the performance study. **a** Patient samples ($n = 50$) and technical triplicates (symbols) with zoom into low concentration range (inset) for renin measured with MR and PR with Passing-Bablok regression (grey line), 95% CI (grey region) and identity line (grey dashed line). **b** same as in **a** for aldosterone, ignoring the limited standard curve fit PR data (dotted line) and additionally data with outliers (solid line, PB: y=0.31x+193.46, slope 95% CI = [0.25, 0.57], intercept 95% CI = [75.62, 259.54]). **c** same as in **a** but MR compared to SOTA (Labcorp). Labcorp values were converted from renin plasma activity (PRA; $\mathrm{ng\,mL^{-1}\,h^{-1}}$) to direct renin concentration (DRC; $\mathrm{pg\,mL^{-1}}$) using the conversion factor 7.6. **d** Same as in **c** for aldosterone; no conversion factor applied. **e,f** same as in **c,d** signals quantified with PR for renin and aldosterone, respectively.

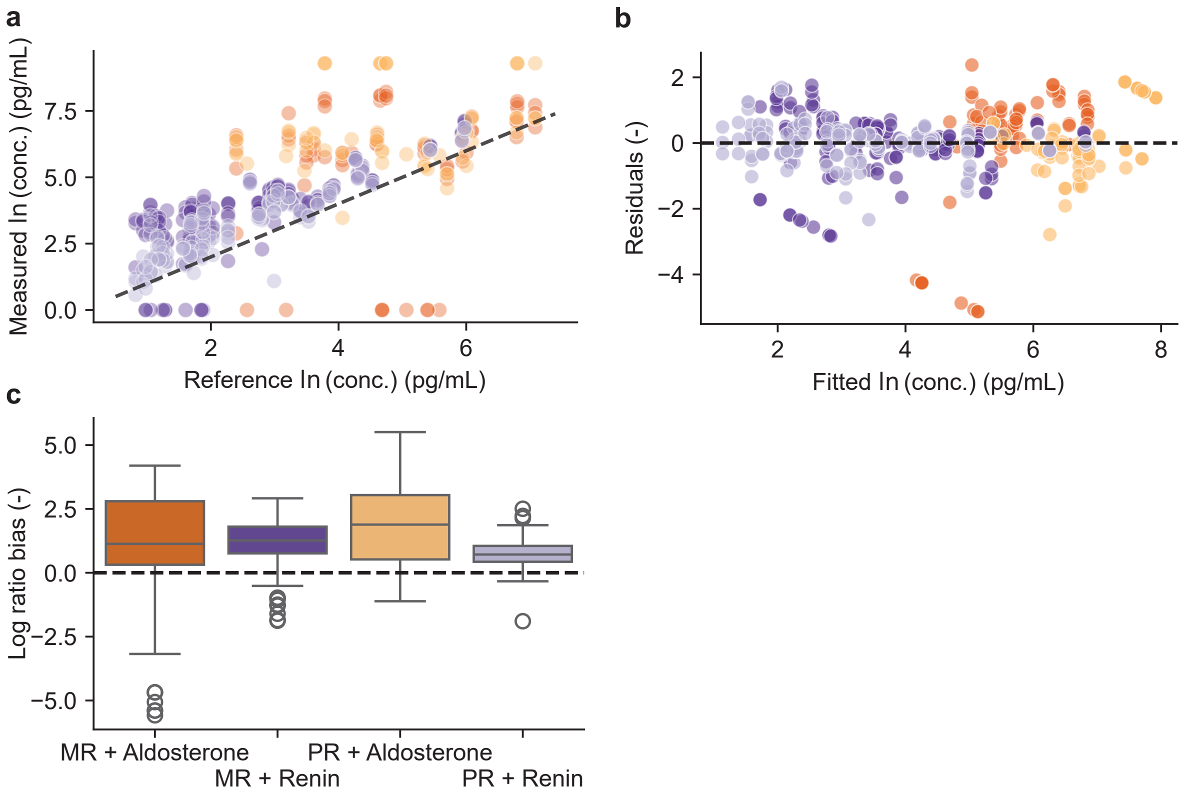


Suppl. Fig. 16: Device performance and calibration relative to LC/MS reference assessed with mixed effect modeling. **a** Scatterplots of MR and PR measurements against Labcorp LC/MS reference. Each color represents a device × assay combination (MR+Aldosterone (orange), MR+Renin (violet), PR+Aldosterone (light orange) and PR+Renin (light violet)). The dashed line indicates perfect agreement (slope = 1). Deviations from the line indicate proportional bias; (n=432). **b** Residuals of the calibration model plotted against fitted log-transformed concentrations by device × assay (coloring same as in **a**. Horizontal line at 0 highlights systematic deviations. Residual structure shows no major device-specific trends, supporting interchangeability **c** Boxplots of log(measured/reference) showing proportional proportional bias for MR and PR across assays. The box represents the inter-quartile range (IQR), with the median shown as a horizontal black line. Whiskers extend to the most extreme data points within 1.5 times the IQR, Outliers (black circles). Values near zero indicate minimal proportional bias; (n=432).

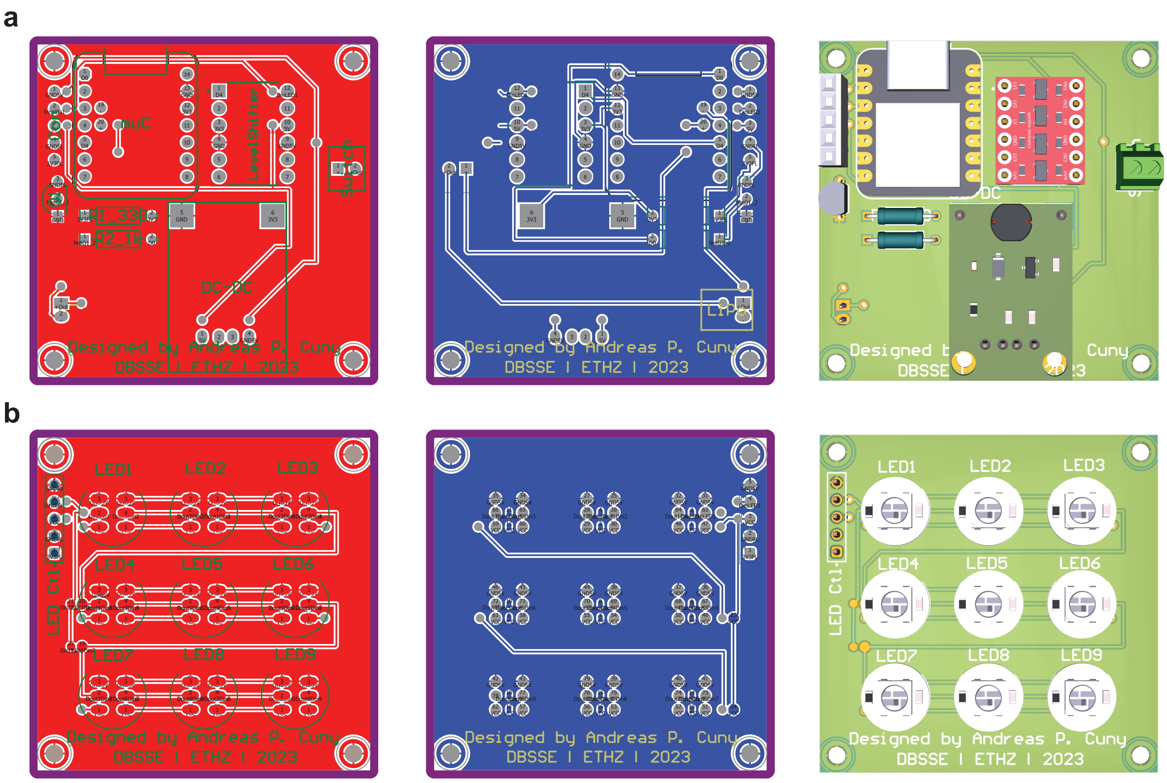


Suppl. Fig. 17: PCB design files for the MCCS. **a** MCCS Base PCB, top (left), bottom layer (right). **b** MCCS LED PCB, top (left), bottom layer (right). Bill of materials (BOM) listed in **Table 11**

Table 1: Camera sensor reliability with signal (mean ± SD) and percent coefficient of variation %CV for the two sensors HP20 and X13P and the region of interest (ROI) and background (BG) metric. The signal was set with the MCCS and ND filters to be just above the noise level.

| Smartphone | Metric | Signal (mean ± SD) | % CoV |
|---|---|---|---|
| HP20 | BG | 8564.13 ± 167.05 | 1.95 |
| HP20 | ROI | 8768.58 ± 158.86 | 1.81 |
| X13P | BG | 6940.27 ± 59.13 | 0.85 |
| X13P | ROI | 7246.66 ± 167.66 | 2.31 |

Table 2: Maximum number of ND filters that could be added and their corresponding theoretical optical power (W) that allowed the distinction between the brightness of a signal from the background for maximum camera settings.

| | ND filters (# x OD) | Optical Power (W) | ISO/Gain | Exposure time (s) |
|---|---|---|---|---|
| X13P | 17 x 0.9 | $7.09 * 10^{-23}$ | 3200 | 30 |
| HP60P | 9 x 0.9 | $1.12 * 10^{-15}$ | 3200 | 30 |
| HP20 | 8 x 0.9 | $8.93 * 10^{-15}$ | 3200 | 30 |
| F5 | 8 x 0.9 | $8.93 * 10^{-15}$ | 3200 | 8 |
| NG22 | 5 x 0.9 | $4.48 * 10^{-12}$ | 9000 | 5 |
| AC3MP | 4 x 0.9 | $3.56 * 10^{-11}$ | 31 | $1.4 * 10^{-3}$ |
| AC5MP | 4 x 0.9 | $3.56 * 10^{-11}$ | 1023 | $30.0 * 10^{-3}$ |
| OV5640 | 3 x 0.9 & 1 x 0.3 | $1.42 * 10^{-10}$ | 63 | $26.0 * 10^{-3}$ |

Table 3: Requested Plasma Renin Activities (PRA) and aldosterone (Aldo.) concentrations of paired human plasma-EDTA and serum samples from Labcorp.

| Results | Sample Type |
|---|---|
| PRA - 3.5 $ng\,mL^{-1}\,h^{-1}$ AND Aldo. - Any $ng\,dL^{-1}$ | Paired serum / plasma-EDTA |
| PRA - 1.7-3.5 $ng\,mL^{-1}\,h^{-1}$ AND Aldo. - Any $ng\,dL^{-1}$ | Paired serum / plasma-EDTA |
| PRA - $<$1.7 $ng\,mL^{-1}\,h^{-1}$ AND Aldo. - Any $ng\,dL^{-1}$ | Paired serum / plasma-EDTA |
| PRA - Any $ng\,mL^{-1}\,h^{-1}$ AND Aldo. - $>$25 $ng\,dL^{-1}$ | Paired serum / plasma-EDTA |
| PRA - Any $ng\,mL^{-1}\,h^{-1}$ AND Aldo. - 10-25 $ng\,dL^{-1}$ | Paired serum / plasma-EDTA |
| PRA - Any $ng\,mL^{-1}\,h^{-1}$ AND Aldo. - $<$10 $ng\,dL^{-1}$ | Paired serum / plasma-EDTA |

Table 4: Different conversion factors and their ranges for Plasma Renin Activity (PRA) to Direct Renin Concentration (DRC) conversion.

| PRA ($ng\,mL^{-1}\,h^{-1}$) | DRC ($pg\,mL^{-1}$) |
|---|---|
| 1 | 7.6 [5.5, 9.7] [1] |
| 1 | [13.69, 18.2] [2, 4] |
| 1 | [20.0, 26.6] [3, 4] |

[1] Warde Lab
[2] 8.2 $mU\,L^{-1}$[1]
[3] 12 $mU\,L^{-1}$[2]
[4] $mU\,L^{-1}$ to $pg\,mL^{-1}$ converted with factors 1.67[3] and 2.22[4]

Table 5: Assay reagents used for aldosterone and renin CLIA listing supplier, catalogue number (Cat. Nr) and Lot number (Lot).

| Reagent | Supplier | Cat. Nr. | Lot |
|---|---|---|---|
| Antibody diluent (HAMA Blocker) for ELISA | abcam | ab193969 | GR3425073-6 |
| LowCross HRP-Stabilizer | Candor Bioscience GmbH | 270500 | 270R248x |
| 10x Phosphate buffer saline (PBS) | Thermo Scientific | J62036.K2 | X26I515 |
| Bovine serum albumin (BSA) | Roth | 0763.3 | 103334024 |
| ProClin300 | Sigma-Aldrich | 48914-U | MKCP7878 |
| Tween-20 | Sigma-Aldrich | P7949-500ML | BCCH4932 |
| Dynabeads M-280 Streptavidin | Invitrogen | 11205D | 2836952 |
| DMSO (dimethylsulfoxide), anhydrous | Invitrogen | D12345 | 250219005 |
| Renin/Prorenin (REN/PREN) Double Depleted Human Plasma | Innovative Research Inc. | IRENPRENDP 10ML | 521 |
| Human Serum | Innovative Research Inc. | ISERBCDA 500ML | 45833 |
| Aldosterone | Sigma-Aldrich | A9477-5MG | MKCP7899 |
| Methanol | Sigma-Aldrich | 34860-2.5L-R | STBK6894 |
| Recombinant human Renin protein | Abcam | ab135012 | GR3445708-3 |
| Luminata™ Crescendo | Merck Millipore | ELLUR0100 | 2317701 |

Table 6: Materials used for reagent preparation and CLIA methods listing supplier, catalogue number (Cat. Nr) and Lot number (Lot).

| Material | Supplier | Cat. Nr. | Lot |
|---|---|---|---|
| Vivaspin® 500 Centrifugal concentrator Membrane 30000 MWCO PES | Sartorius | VS0122 | 235000067 |
| Sterile Filter Unit 0.22$\mu m$ | Merck Millipore | SLGS033SB | 0000429304 |
| EZ-Link™ Plus Activated Peroxidase Ki | Thermo Scientific | 31487 | PF202431A |
| EZ-Link NHS-PEG4-Biotin | Thermo Scientific | 21362 | UH287569 |
| Costar Assay plate 96-well (high binding, clear) | greiner | 655101 | E16103QM |
| C8 Maxisorp white | Thermo Scientific | 463201 | 175641 |
| SealPlate | EXCEL Scientific | 100SEALPLT | SH332A |

Table 7: Affinity reagents used in aldosterone and renin CLIAs.

| Reagent | Supplier | Cat. Nr. | Lot |
|---|---|---|---|
| Aldosterone-3-CMO-HRP-Enzyme-Conjugate | Pantex | n/a | ALDEC-103 |
| Anti-h Renin 10850 | Medix Biochemica | 700025 | 2004482 |
| Anti-h Renin 10852 | Medix Biochemica | 700030 | 2004100 |
| Anti-aldosterone HM783 | Medix Biochemica | HM783 | B2628 |

Table 8: Limits of the standard curves. Blank (mean and standard deviation, SD), percent coefficient of variation of blank, standard deviation of the lowest non-zero sample (LNZ, SD), lower limit of detection (LLoD), limit of quantification (LoQ), and upper limit of detection (ULoD). Limits were calculated following FDA/ICH guidelines and Armbruster et cal.[5–8] as $\text{LoB} = \mu_b \pm 1.645\,\sigma_b$, $\text{LoD} = \mu_b \pm 3.3\,\sigma_b$, $\text{LoQ} = \mu_b \pm 10\,\sigma_b$, $\text{LoD}_{\text{blank}} = \text{LoB} \pm 1.645\,\sigma_{LNZ}$, and $\text{ULoD} = \mu_h \pm 1.645\,\sigma_h$, with mean blank $\mu_b$, SD blank $\sigma_b$, SD LNZ $\sigma_{LNZ}$, mean highest non saturated concentration $\mu_h$ and its SD $\sigma_h$ where the sign depends on the assay type (increasing or decreasing signal with concentration). Note these are preliminary limits and may change during further development of the assays and reaching recommended 20-60 technical replicates.

| Figure | Reader | Processing | Assay | Blank (mean ± SD) | %CV (Blank) | $SD_{LNZ}$ | LLoD (pg mL$^{-1}$) | LLoDB[1] (pg mL$^{-1}$) | LoQ (pg mL$^{-1}$) | ULoD (pg mL$^{-1}$) |
|---|---|---|---|---|---|---|---|---|---|---|
| **Fig. 4f** | MR | RAW | renin | 5472.81 ± 158.43 | 2.89 | 147.63 | 13.16 | 12.80 | 30.46 | 871.39 |
| **Fig. 4f** | MR | ROF | renin | 5490.40 ± 144.26 | 2.63 | 131.86 | 12.37 | 11.95 | 28.49 | 870.85 |
| **Fig. 4f** | MR | $\text{NREA}_{\text{ROF}}$ | renin | 4252.13 ± 53.05 | 1.25 | 105.37 | 1.53 | 2.07 | 3.54 | 717.88 |
| **Fig. 4g** | MR | RAW | aldosterone | 51201.28 ± 981.23 | 1.92 | 887.01 | 26.91 | 26.76 | 33.84 | 1580.01 |
| **Fig. 4g** | MR | ROF | aldosterone | 51154.90 ± 993.49 | 1.94 | 889.58 | 26.96 | 26.79 | 34.06 | 1567.12 |
| **Fig. 4g** | MR | $\text{NREA}_{\text{ROF}}$ | aldosterone | 239450.68 ± 10087.45 | 4.21 | 10692.45 | 30.79 | 30.97 | 53.08 | 1805.83 |
| **Fig. 5a** | MR | $\text{NREA}_{\text{ROF}}$ | renin | 4252.13 ± 53.05 | 1.25 | 105.37 | 1.53 | 2.07 | 3.54 | 717.88 |
| **Fig. 5b** | MR | $\text{NREA}_{\text{ROF}}$ | aldosterone | 239450.68 ± 10087.45 | 4.21 | 10692.45 | 30.79 | 30.97 | 53.08 | 1805.83 |
| **Suppl. Fig. 11a** | PR | RAW | renin | 889.50 ± 433.46 | 48.73 | 17.68 | 4.01 | 3.09 | 8.10 | 9769.66 |
| **Suppl. Fig. 11b** | PR | RAW | aldosterone | 455021.50 ± 11133.40 | 2.45 | 2279.01 | 12.64 | 5.26 | 32.06 | 1682.25 |
| **Fig. 6a** | MR | $\text{NREA}_{\text{ROF}}$ | renin | 7823.72 ± 509.62 | 6.51 | 592.98 | 27.93 | 29.62 | 62.79 | 2604.48 |
| **Suppl. Fig. 13a** | MR | $\text{NREA}_{\text{ROF}}$ | aldosterone | 269114.46 ± 15254.44 | 5.67 | 16665.00 | 879.89 | 918.01 | 2806.11 | 1460.31 |
| **Suppl. Fig. 14b** | PR | RAW | renin | 154.17 ± 65.72 | 42.63 | 105.42 | 4.28 | 5.00 | 8.84 | 4719.60 |
| **Suppl. Fig. 14e** | PR | RAW | aldosterone | 244000.17 ± 15900.85 | 6.52 | 22905.06 | 871.94 | 1054.61 | 10855.03 | 1310.25 |

[1] More conservative estimation of the limit of detection according to[8].

Table 9: Table of Correlations with the $R^2$, Pearson correlation $r$ [95% CI], $p-Value$, Standard error (Std. Error) and number of datapoints ($N$).

| Figure | Reader | Assay | Correlation | $R^2$ | $r$ [95% CI] | $p-Value$ | Std. Error | $N$ |
|---|---|---|---|---|---|---|---|---|
| **Fig. 5c** | MR | renin | MR-PR | 0.994 | 1 [0.99, 1] | 2.23e-12 | 0.02 | 12 |
| **Fig. 5d** | MR | aldosterone | MR-PR | 0.906 | 0.95 [0.83, 0.99] | 1.84e-06 | 0.06 | 12 |
| **Fig. 5e** | MR | renin | MR-Labcorp | 0.869 | 0.93 [0.77, 0.98] | 9.98e-06 | 0.11 | 12 |
| **Fig. 5f** | MR | aldosterone | MR-Labcorp | 0.544 | 0.74 [0.28, 0.92] | 6.19e-03 | 0.02 | 12 |
| **Suppl. Fig. 11c** | PR | renin | PR-Labcorp | 0.913 | 0.96 [0.85, 0.99] | 1.27e-06 | 0.09 | 12 |
| **Suppl. Fig. 11d** | PR | aldosterone | PR-Labcorp | 0.514 | 0.72 [0.24, 0.91] | 8.73e-03 | 0.03 | 12 |
| **Fig. 6b** | MR | renin | MR-PR | 0.990 | 1 [0.99, 1] | 3.57e-151 | 0.01 | 150 |
| **Fig. 6c** | MR | renin | MR-Labcorp | 0.934 | 0.97 [0.95, 0.98] | 3.25e-89 | 0.05 | 150 |
| **Suppl. Fig. 14c** | PR | renin | PR-Labcorp | 0.931 | 0.96 [0.95, 0.97] | 1.04e-87 | 0.04 | 150 |
| **Suppl. Fig. 13b** | MR | aldosterone | MR-PR | 0.812 | 0.9 [0.84, 0.94] | 6.27e-25 | 0.01 | 66 |
| **Suppl. Fig. 13b** | MR | aldosterone | MR-$PR_{sub}$ [1] | 0.514 | 0.72 [0.55, 0.83] | 1.56e-09 | 0.12 | 53 |
| **Suppl. Fig. 13c** | MR | aldosterone | MR-Labcorp | 0.056 | 0.24 [-0.01, 0.45] | 5.64e-02 | 0.38 | 66 |
| **Suppl. Fig. 13d** | PR | aldosterone | PR-Labcorp | 0.036 | 0.19 [-0.05, 0.41] | 1.26e-01 | 1.57 | 66 |

[1] Excluding PR data outside stc model fit.

Table 10: Bias, calibration slopes, proportional bias significance, and sample-level ICC for device × assay combinations.

| Planned Comparison | Bias Estimate [95% CI] | P-value | Calibration slope [95% CI] | Slope P vs 1 | ICC [sample] |
|---|---|---|---|---|---|
| MR (Aldosterone) vs. 0 | 598.72 [179.71, 1017.72] | 0.005 | 0.40 [0.18, 0.62] | 1.65e-07 | 0.21 |
| PR (Aldosterone) vs. 0 | 2261.53 [1842.52, 2680.54] | 3.74e-26 | 0.29 [0.06, 0.51] | 5.16e-10 | 0.21 |
| MR (Renin) vs. 0 | 65.79 [-239.79, 371.37] | 0.673 | 0.83 [0.66, 1.00] | 0.0436 | 0.21 |
| PR (Renin) vs. 0 | 41.82 [-263.77, 347.40] | 0.789 | 0.95 [0.79, 1.12] | 0.575 | 0.21 |
| PR vs. MR (Aldosterone) | 1662.81 [1176.49, 2149.14] | 2.06e-11 | - | - | 0.21 |
| PR vs. MR (Renin) | -23.97 [-346.57, 298.62] | 0.884 | - | - | 0.21 |

Table 11: MCCS Bill of Materials

| Ref | Qnty | Value | Footprint [1] | Price (10 units) | Total Cost | Digikey Link |
|---|---|---|---|---|---|---|
| **MCCS LED Board** | | | | | | |
| LED1-9 | 9 | Adafruit Industries 1612 | C_0603_1608Metric | $ 0.21 | $ 1.85 | 1528-1196-ND |
| LED Ctl | 1 | A106751-ND | CONN HEADER VERT 5POS | $ 0.53 | $ 0.53 | A106751-ND |
| | | | | Subtotal: | $ 2.37 | |
| **MCCS BASE Board** | | | | | | |
| muC | 1 | XIAO NRF52840 | 102010448 | $9.96 | $ 9.96 | 1597-102010448-ND |
| DC-DC | 1 | DIYMORE DC-DC 5V, 500mA | 2 layer board | $3.99 | $ 3.99 | N/A [2] |
| LevelShift | 1 | SparkFun 4-CH level shifter | BOB-12009 | $3.95 | $ 3.95 | 1568-12009-ND |
| Q1 | 1 | ON Semiconductor PN2222ATA | PN2222ATA | $0.19 | $ 0.19 | PN2222ATATB-ND |
| R1_330 | 1 | 330Ohm | | $0.13 | $ 0.13 | A138238TB-ND |
| R2_1k | 1 | 1kOhm | | $0.13 | $ 0.13 | A138131TB-ND |
| Header | 1 | A32906-ND | Connector PinHeader 2.54mm:PinHeader 1x05 P2.54mm | $ 1.70 | $ 1.70 | A32906-ND |
| Switch | 1 | TE Connectivity 282834-2 | | $1.35 | $ 1.35 | A98333-ND |
| LiPo | 1 | JST S2B-PH-K-S (LF)(SN) | | $0.10 | $ 0.10 | 455-1719-ND |
| Battery | 1 | LiPo Battery Pack | N/A, 37x30.5x5.3mm | $7.95 | $ 7.95 | 1528-1841-ND |
| Rocker Switch | 1 | Rocker Switch SPST | N/A, 14.40mm Dia | $0.39 | $ 0.39 | 2057-SW-R3-1A-A-1-2-ND |
| | | | | Subtotal: | $ 29.83 | |
| **PCB Fab** | | | | | | |
| MCCS LED PCB | 1 | MCCS LED Shield | 2 layer board | 1 | $ 1.00 | |
| MCCS BASE PCB | 1 | MCCS Base Board | 2 layer board | 1 | $ 1.00 | |
| | | | | Total cost: | $ 34.20 | |

[1] Digikey provides EDA/CAD models in different formats for common PCB design software.
[2] Different supplier.

Table 12: Table of 5PL fitting parameters used in the figures, for a given assay, reader and processing. The 5PL model parameters [with 95% CI], a (lower asymptote), b (slope), c (inflection point), d (upper asymptote), and g (asymmetry factor). R-square ($R^2$), and delta corrected Akaike information criterion (ΔAICc)

| Figure | Reader | Processing | Assay | a [95% CI] | b [95% CI] | c [95% CI] | d [95% CI] | g [95% CI] | $R^2$ | ΔAICc |
|---|---|---|---|---|---|---|---|---|---|---|
| **Fig. 4f** | MR | RAW | renin | 5.5e+03 [5.4e+03, 5.5e+03] | 1.4 [1.3, 1.5] | 1.5e+02 [1.3e+02, 1.8e+02] | 1.1e+05 [9.7e+04, 1.2e+05] | 0.15 [0.11, 0.19] | 0.99 | 0.00 |
| **Fig. 4f** | MR | ROF | renin | 5.5e+03 [5.4e+03, 5.5e+03] | 1.4 [1.3, 1.5] | 1.5e+02 [1.3e+02, 1.7e+02] | 1.1e+05 [9.8e+04, 1.2e+05] | 0.14 [0.1, 0.18] | 0.99 | 0.00 |
| **Fig. 4f** | MR | $NREA_{ROF}$ | renin | 4.3e+03 [4.2e+03, 4.3e+03] | 1.3 [1.2, 1.4] | 1.6e+02 [60, 2.7e+02] | 6.3e+05 [3.8e+04, 1.2e+06] | 0.14 [-0.082, 0.36] | 0.99 | 0.00 |
| **Fig. 4g** | MR | RAW | aldosterone | 5.1e+04 [5.1e+04, 5.1e+04] | 26 [-1.8e+04, 1.8e+04] | 25 [19, 30] | 5.8e+03 [5.2e+03, 6.4e+03] | 0.029 [-20, 20] | 0.99 | 0.00 |
| **Fig. 4g** | MR | ROF | aldosterone | 5.1e+04 [5.1e+04, 5.1e+04] | 24 [-1e+04, 1e+04] | 25 [19, 30] | 5.8e+03 [5.2e+03, 6.4e+03] | 0.031 [-13, 13] | 0.99 | 0.00 |
| **Fig. 4g** | MR | $NREA_{ROF}$ | aldosterone | 2.4e+05 [2.3e+05, 2.5e+05] | 26 [-4.5e+04, 4.5e+04] | 25 [2.6, 47] | 7.8e+03 [-5.2e+03, 2.1e+04] | 0.030 [-52, 52] | 0.99 | 2.62[1] |
| **Fig. 5a** | MR | $NREA_{ROF}$ | renin | 4.3e+03 [4.2e+03, 4.3e+03] | 1.3 [1.2, 1.4] | 1.6e+02 [60, 2.7e+02] | 6.3e+05 [3.8e+04, 1.2e+06] | 0.14 [-0.082, 0.36] | 0.99 | 0.00 |
| **Fig. 5b** | MR | $NREA_{ROF}$ | aldosterone | 2.4e+05 [2.3e+05, 2.5e+05] | 26 [-4.5e+04, 4.5e+04] | 25 [2.6, 47] | 7.8e+03 [-5.2e+03, 2.1e+04] | 0.030 [-52, 52] | 0.99 | 2.62[1] |
| **Suppl. Fig. 11a** | PR | | renin | -1e+03 [-1.5e+03, -4.7e+02] | 0.9 [0.87, 0.93] | 2.1e+03 [1.6e+03, 2.7e+03] | 2.7e+08 [2.7e+08, 2.7e+08] | 0.0034 [0.0029, 0.0039] | 0.97 | 0.00 |
| **Suppl. Fig. 11b** | PR | | aldosterone | 4.4e+05 [4.3e+05, 4.4e+05] | 2.1 [0.89, 3.3] | 41 [8.7, 72] | 2.2e+04 [1.2e+04, 3.1e+04] | 0.53 [-0.096, 1.2] | 0.99 | 8.21[1] |
| **Fig. 6a** | MR | $NREA_{ROF}$ | renin | 8.1e+03 [7.8e+03, 8.3e+03] | 1.5 [1.4, 1.6] | 9.2e+03 [-6.5e+04, 8.4e+04] | 3.3e+05 [3.2e+05, 3.3e+05] | 30 [-3.3e+02, 3.9e+02] | 0.99 | 0.00 |
| **Suppl. Fig. 13a** | MR | $NREA_{ROF}$ | aldosterone | 2.7e+05 [2.6e+05, 2.9e+05] | 0.9 [0.0085, 1.8] | 7.1e+06 [-3.0e+10, 3.0e+10] | -2.0e+07 [-1.1e+10, 1.1e+10] | 9 [-3.6e+04, 3.6e+04] | 0.74 | 2.39[1] |
| **Suppl. Fig. 14b** | PR | | renin | 62 [16, 1.1e+02] | 1.2 [1.2, 1.2] | 2.6e+03 [2.2e+03, 3e+03] | 1.1e+08 [1.1e+08, 1.1e+08] | 7.2e-03 [6.2e-03, 8.1e-03] | 0.99 | 5.06[1] |
| **Suppl. Fig. 14e** | PR | | aldosterone | 2.4e+05 [2.3e+05, 2.5e+05] | 1.6 [-0.94, 4.2] | 9.1e+03 [-1.7e+06, 1.8e+06] | 1.4e+05 [-1.2e+05, 3.9e+05] | 30 [-9.3e+03, 9.3e+03] | 0.84 | 0.00 |

[1] to 4PL
